# Genetic liability to metabolic dysfunction modelled in early adulthood predicts cardiometabolic risk across the life course in Asian populations

**DOI:** 10.64898/2026.04.24.26351660

**Authors:** Hong Pan, Evelyn Lau, Yurui Chen, Jian Huang, Ai Ling Teh, Pei Fang Tan, Shaun Seh Ern Loong, Kazuo Miyazawa, Xiaohe Zhang, Varsha Gupta, Priti Mishra, Jinyi Che, Nicholas Li, Eran Elhaik, Suresh Anand Sadananthan, Sambasivam Sendhil Velan, Kuang Lin, Robin G. Walters, Juan A. Delgado-SanMartin, Kok Hian Tan, Jerry Kok Yen Chan, Carolyn SP Lam, Arthur Mark Richards, Koichi Matsuda, Kaoru Ito, Roger Foo, Johan Gunnar Eriksson, Karen Mei Ling Tan, David Cameron-Smith, Dennis Wang

**Affiliations:** Institute for Human Development and Potential (IHDP), Agency for Science, Technology and Research (A*STAR), Singapore, Republic of Singapore; Bioinformatics Institute (BII), Agency for Science, Technology and Research (A*STAR), Singapore, Republic of Singapore; Department of Mathematics and Computational Biology Programme, National University of Singapore, Republic of Singapore; Yong Loo Lin School of Medicine, National University of Singapore, Republic of Singapore; Cardiovascular Disease Translational Research Program Institute, National University of Singapore, Republic of Singapore; Laboratory for Cardiovascular Genomics and Informatics, RIKEN Center for Integrative Medical Sciences, Yokohama, Japan; Department of Biology, Lund University, Lund, Sweden; Nuffield Department of Population Health, University of Oxford, Oxford, United Kingdom; National Heart & Lung Institute, Imperial College London, London, United Kingdom; Academic Clinical Program in Obstetrics and Gynaecology, Duke- National University of Singapore Medical School, Republic of Singapore; Department of Maternal Fetal Medicine, KK Women’s and Children’s Hospital, Republic of Singapore; Department of Reproductive Medicine, KK Women’s and Children’s Hospital, Republic of Singapore; National Heart Centre Singapore, Republic of Singapore; Duke-National University of Singapore Medical School, Republic of Singapore; Baim Institute for Clinical Research, Boston MA, USA; Cardiovascular Research Institute, National University of Singapore, Republic of Singapore; Christchurch Heart Institute, University of Otago, New Zealand; Institute of Medical Science, The University of Tokyo, Laboratory of Genome Technology, Human Genome Center, Tokyo, Japan; Department of Computational Biology and Medical Sciences, Laboratory of Clinical Genome Sequencing, Graduate School of Frontier Sciences, The University of Tokyo, Tokyo, Japan; Department of Laboratory Medicine, National University Hospital, Republic of Singapore; Riddet Institute, Massey University, Palmerston North, New Zealand

**Keywords:** Metabolic syndrome, Cardiometabolic disease, Multimorbidity, Polygenic risk score, Asian populations, Life course, Genetic susceptibility, Population health

## Abstract

**Background:** Cardiometabolic diseases arise from metabolic dysfunction that develops decades before clinical onset. Conventional genetic risk models are typically derived in middle-aged or older populations, where genetic effects are confounded by cumulative environmental exposures, chronic comorbidities, and clinical interventions. Whether the life stage at which genetic liability is modelled influences the biological signal captured by polygenic scores remains unclear, particularly in underrepresented populations. We therefore tested whether genetic liability modelled in early adulthood, a period of relative physiological stability, is associated with cardiometabolic risk across the life course in Asian populations.

**Methods:** We developed a polygenic score for metabolic syndrome, GenMetS, using data from 1,368 Singaporean women aged 18–45 years. The model integrates 15 established polygenic scores for metabolic traits and applies elastic-net penalized regression to optimize variant weights. GenMetS was evaluated in five cohorts comprising 670,952 individuals aged 0–94 years across population-based and disease-enriched settings, including Asian and European ancestry groups. Associations with metabolic traits, cardiometabolic diseases, multimorbidity, and early-life growth patterns were assessed.

**Results:** In Asian populations, GenMetS explained 5.0–12.4% of the variance in metabolic syndrome in adults and 10.3% in children, with negligible performance in European populations (R² < 0.001). Higher GenMetS was associated with increased odds of cardiometabolic diseases, including type 2 diabetes, heart failure, and stroke (odds ratios 1.32–1.52 per standard deviation). In UK Biobank participants of Asian ancestry, GenMetS improved discrimination of cardiometabolic multimorbidity beyond age alone. Associations were consistent across sexes. In children, higher GenMetS was associated with obesogenic growth trajectories and increased abdominal adiposity.

**Conclusions:** Genetic liability to metabolic dysfunction modelled in early adulthood captures a stable biological signal associated with metabolic traits, disease risk, and multimorbidity from childhood to adulthood in Asian populations. These findings indicate that the life stage of model derivation shapes the biological signal captured by polygenic scores and support the development of life-stage- and ancestry-informed approaches for cardiometabolic risk assessment and prevention.

## Background

Metabolic syndrome reflects the coordinated dysregulation of adiposity, glucose metabolism, lipid homeostasis, and vascular regulation, and represents an early manifestation of cardiometabolic liability that often precedes the clinical onset of type 2 diabetes (T2D) and cardiovascular disease by several decades [1–5]. The burden of these conditions is rising rapidly across Asian populations [6–8], where metabolic complications frequently emerge at younger ages and at higher levels of body fat despite lower body mass index (BMI) than in European populations [9–11]. These features indicate a distinct pattern of metabolic risk in Asian populations and underscore the need for ancestry-informed models to accurately characterize underlying risk.

Despite this need, polygenic scores (PGS) for metabolic syndrome and its component traits have been derived predominantly from European cohorts [12–14]. Their performance is lower in Asian populations, as differences in allele frequencies and linkage disequilibrium structure limit cross-ancestry applicability [15–17]. Furthermore, most PGS are also trained using metabolic traits measured in middle-aged or older adults [12, 14]. In these settings, genetic effects are difficult to separate from cumulative environmental exposures, chronic comorbidities, and medication use [18]. Consequently, it remains unclear whether current models capture underlying genetic liability or reflect downstream clinical and treatment-related effects. It is also unclear whether the life stage at which polygenic scores are constructed affects the biological signal they represent.

During early to mid-adulthood, metabolic physiology is relatively stable and the prevalence of chronic disease and medication use is low, allowing genetic effects to be evaluated with minimal influence from clinical confounding [19–22]. This stage may therefore provide a clearer window to capture genetic influences on metabolic regulation. In Asian women, early metabolic perturbations during this period are particularly consequential [20], as these perturbations often track into later-life cardiometabolic disease and intersect with female-specific conditions such as gestational diabetes, a strong predictor of future T2D and cardiovascular risk [23–25]. Although these perturbations are particularly relevant in this demographic, the autosomal variants underlying core metabolic pathways are shared across sexes, supporting the evaluation of genetic liability in both women and men.

In this study, we developed GenMetS, a polygenic score trained in 1,368 young adult women in Singapore, to model genetic liability during early adulthood, a stage with reduced clinical and treatment-related confounding. We evaluated GenMetS in 670,952 individuals across five cohorts spanning birth to 94 years of age, including population-based and disease-enriched settings across Asia and Europe. We assessed its associations with metabolic traits, cardiometabolic diseases, multimorbidity, and early-life growth patterns to test the hypothesis that genetic liability modelled during early adulthood captures a signal that remains associated with cardiometabolic outcomes across the life course. As the model is based on autosomal variants underlying core metabolic pathways, cross-sex evaluation is biologically plausible.

## Methods

### Study design and cohorts

The GenMetS model was developed using 1,368 young adult women of Asian ancestry aged 18-45 years from two prospective cohorts, the Singapore Preconception Study of Long-Term Maternal and Child Outcomes [21] (S-PRESTO; n=934) and Growing Up in Singapore Towards Healthy Outcomes (GUSTO; n=434) [19]. The discovery cohort included 934 women from S-PRESTO assessed prior to pregnancy and 434 women from GUSTO assessed at 8 years postpartum (**Table S1**). In total, 1,368 participants with complete genotype data, metabolic measurements, and a composite metabolic syndrome score were included. The mean age was 33.98 years (standard deviation (SD), 5.72). Participants self-identified as Chinese, Malay, Indian, or of mixed Asian ancestry.

The performance of GenMetS was evaluated across five cohorts spanning the life course from birth to 94 years of age (**Figure 1**). Validation included 365,495 participants from the UK Biobank [26] of European and Asian ancestry, with primary disease and multimorbidity analyses conducted in the Asian-ancestry subset (n=8,792) to ensure ancestry concordance with the discovery phase. The UK Biobank cohort comprised 195,665 women at mean (SD) age 56.60 (7.94) years and 169,830 men at 56.98 (8.13) years (**Table S1 and Fig. S1**). Clinical performance in high-risk settings was assessed in the ATTRaCT cohort (The Asian neTwork for Translational Research and Cardiovascular Trials), comprising 2,310 Asian adults enriched for heart failure cases [27, 28], with a mean age of 58.78 (10.45) years in women and 58.48 (10.43) years in men. Further validation was performed in the Biobank Japan, a hospital-based national biobank of approximately 200,000 patients with a mean age of 64 years [29, 30] (**Table S2**), and in the China Kadoorie Biobank [31], where genotype and disease outcomes from 100,706 participants were used for GenMetS evaluation [32].

**Fig. 1.**
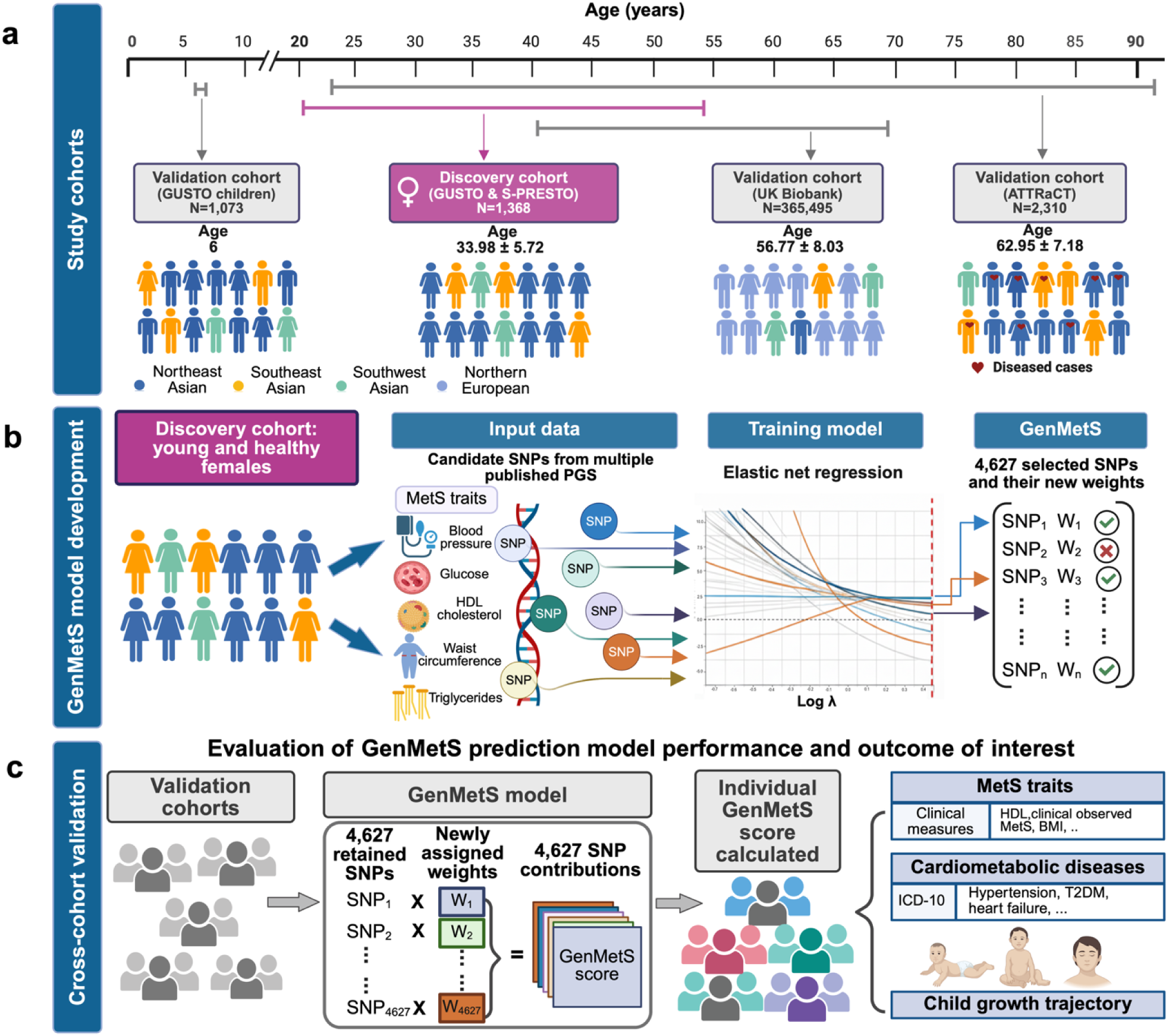
Overview of study cohorts, GenMetS model development, and cross-cohort evaluation. **a,** Study cohorts included in model development and primary validation. The GenMetS model was developed in a discovery cohort comprising 1,368 Asian women aged 18–45 years from GUSTO and S-PRESTO. The model was then validated in participants from multi-ethnic cohorts spanning childhood to older adulthood. Validation cohorts included GUSTO children (N = 1,073; ages 0-6 years), UK Biobank adults (N = 365,495; predominantly European with a minority Asian subset), and ATTRaCT adults (N = 2,310; cardiometabolic disease cohort). Major genetic ancestry groups represented in these cohorts include Northeast Asian, Southeast Asian, Southwest Asian, and Northern European. Biobank Japan and the China Kadoorie Biobank were used for disease association analyses. **b,** GenMetS model development. Input data were from the discovery cohort, with the metabolic syndrome (MetS) score measured from its component traits (waist circumference, triglycerides, fasting glucose/HbA1c, blood pressure, and HDL cholesterol) and the genotype data for candidate SNPs aggregated from publicly available polygenic scores associated with metabolic traits. Elastic-net regression was applied to estimate SNP weights associated with the MetS score. The final GenMetS model comprised 4,627 SNPs with non-zero weights. **c,** Cross-cohort validation and downstream analyses for GenMetS. GenMetS scores were evaluated in independent validation cohorts to assess predictive performance for: (i) MetS and related metabolic traits, (ii) cardiometabolic diseases including hypertension, type 2 diabetes, myocardial infarction, heart failure, and multimorbidity, and (iii) childhood growth trajectories. Image was created with BioRender.

Longitudinal evaluation was conducted in 1,073 children from the GUSTO cohort who are the biological offspring of the women in the discovery cohort. This paediatric assessment included child genotype data and longitudinal growth measurements from birth to age six, with 500 children receiving deep metabolic phenotyping at the six-year follow-up (**Table S3**). Across all cohorts, genotype generation, imputation, and quality control procedures were standardized, with platform details and procedures documented in the **Supplementary Information**. Ethical oversight was obtained from relevant institutional review boards, and all participants provided written informed consent.

### Genetic ancestry inference

Genetic ancestry was inferred across all cohorts using between 40,000 and 130,000 autosomal ancestry-informative markers shared between datasets [33]. Admixture proportions were estimated through supervised ADMIXTURE, utilizing nine reference ancestral populations to project individuals onto global gene pools. Following this projection, k-means clustering was applied to identify four genetically homogeneous subcontinental clusters: Northeast Asian, Southeast Asian, Southwest Asian, and Northern European (**Table S4)**.

For primary downstream analyses, participants were aggregated into two broad continental categories to ensure consistent comparisons and maximize statistical power: Asian ancestry, comprising the Northeast, Southeast, and Southwest groups, and European ancestry, consisting of the Northern European group. Ancestry distributions were visualized using the R package pophelper (v2.3.1) [34], with detailed cluster profiles and subgroup distributions documented in the **Supplementary Information**.

### Metabolic syndrome and metabolic traits

Metabolic syndrome was defined according to the National Cholesterol Education Program Adult Treatment Panel III criteria [2] as the presence of three or more of the following components: elevated waist circumference, hypertriglyceridemia, low HDL cholesterol, hyperglycemia, or hypertension. Each abnormal component was coded as a binary variable and summed to derive a MetS score ranging from 0 to 5. To stabilize variance for genetic modeling in the discovery cohort, where fewer than 5% of participants exhibited four or more abnormal components, MetS scores of 3 or higher were truncated to 3. MetS scores were used as the outcome for genetic modelling and are summarised across cohorts and age groups in **Table S5**.

Cohort-specific adaptations were implemented to harmonize phenotypes across the validation studies where primary measurements were unavailable. In the ATTRaCT cohort, a body mass index of 25 kg/m² or higher served as a proxy for central adiposity [35]. The validity of this proxy was confirmed through a sensitivity analysis in 81 participants with both measurements available, which demonstrated 70% concordance in MetS classification. For the UK Biobank, where blood samples were predominantly non-fasting, glycated hemoglobin of 5.7% or higher was utilized to define glycemic status [36, 37]. Medication data, including the use of antihypertensive, glucose-lowering, and lipid-modifying agents, were retrieved and incorporated into the component definitions using Anatomical Therapeutic Chemical codes [38].

In the GUSTO offspring cohort, longitudinal anthropometric measurements, including weight, height, and abdominal circumference, were obtained from birth through 72 months. At the 6-year follow-up, fasting blood samples were collected to measure glucose, insulin, and lipid profiles. A pediatric metabolic syndrome score was subsequently constructed using a validated composite formula that integrated age- and sex-standardized z-scores for central adiposity, blood pressure, insulin resistance (HOMA-IR), and lipids [39]. Detailed procedures regarding phenotype harmonization and childhood magnetic resonance imaging are provided in the **Supplementary Information.**

### GenMetS model development

The GenMetS polygenic model was constructed using a two-stage framework that integrated feature aggregation from existing polygenic scores with regularized reweighting in the discovery cohort. In the first stage, candidate genetic variants were compiled from 15 previously published polygenic scores for individual components of metabolic syndrome, including waist circumference, low-density lipoprotein cholesterol, high-density lipoprotein cholesterol, triglycerides, fasting glucose, glycated hemoglobin, and both systolic and diastolic blood pressure. These scores were selected from the PGS Catalog [40] based on trait relevance, sample size, and availability (**Table S6**). Across these studies, a total of 84,129 unique single-nucleotide polymorphisms (SNPs) were identified.

To refine these candidate variants for the discovery cohort, SNPs were pruned for linkage disequilibrium using PLINK (--indep 200 50 0.25), to reduce redundancy, resulting in 31,409 independent markers being retained for model fitting. Metabolic syndrome severity was modelled as a continuous score ranging from 0 to 3, with truncation applied at three components due to sparse observations beyond this threshold in the young adult discovery population.

In the second stage, elastic-net penalized regression was applied to the discovery cohort data to jointly select and reweight these variants. This approach was chosen to facilitate feature selection while retaining correlated SNPs arising from overlapping metabolic pathways, which is appropriate for a multicomponent trait such as MetS. Model hyperparameters were optimized through ten-fold cross-validation. The final GenMetS model retained 4,627 SNPs with non-zero coefficients (**Table S7**). Functional mapping and gene-set enrichment analyses were subsequently conducted using FUMA and MSigDB to characterize the biological processes represented by these variants.

### Association studies and disease prediction

The performance of GenMetS was evaluated through several distinct association analyses across the validation cohorts. The proportion of variance explained in continuous metabolic traits was assessed using linear regression models, with results reported as the coefficient of determination (R^2^). Confidence intervals for these estimates were derived using percentile bootstrap resampling with 1,000 iterations.

For clinical outcomes, we analyzed six major cardiometabolic diseases: type 2 diabetes, hypertension, coronary artery disease, myocardial infarction, heart failure, and stroke. Disease status was ascertained via established clinical coding systems, such as ICD-10 [26], reported in **Table S8**. To evaluate associations, logistic regression models were fitted using the standardized GenMetS score as the primary predictor. Case-control groups were constructed using age-balanced propensity score matching where individual-level data were available, particularly in the UK Biobank and ATTRaCT cohorts. In Biobank Japan and the China Kadoorie Biobank, cohort-specific investigators performed analyses using age-balanced case-control designs and established disease definitions.

To assess the consistency and magnitude of genetic risk across different populations, results were synthesized using fixed-effect inverse-variance weighted meta-analyses. These meta-analyses were conducted separately for women and men, as well as in pooled analyses, to evaluate sex-specific and overall associations. Results are reported as odds ratios (OR) with corresponding 95% confidence intervals per one-standard-deviation (1 SD) increase in genetic risk.

Cardiometabolic multimorbidity, defined as the presence of two or more of the analyzed diseases, was evaluated in UK Biobank participants of Asian ancestry. We implemented extreme gradient boosting (XGBoost) models to compare the predictive utility of GenMetS against five demographic and lifestyle factors: education, socioeconomic status, television viewing time, physical activity, and sleep duration. Specific UK Biobank summary statistics for these variables and disease diagnoses are reported in **Table S9 and Table S10**. These sex-stratified models utilized balanced datasets and were evaluated across 100 bootstrap iterations to ensure robust discrimination, quantified by the area under the receiver operating characteristic curve (AUC). Predictor contributions were further interpreted using SHapley Additive exPlanations (SHAP).

### Statistical analysis and software

GenMetS scores were calculated in all validation cohorts by aligning genotype data to the set of variants and weights derived in the discovery cohort. Scores were computed as the weighted sum of effect allele dosages under an additive genetic model using PLINK (v2.0) (--score with cols=scoreavgs). To ensure comparability across diverse cohorts and age groups, scores were standardized within each cohort to a mean of zero and unit variance, enabling the interpretation of all associations per one standard deviation increase in genetic risk.

For cardiometabolic disease outcomes, including type 2 diabetes, hypertension, coronary artery disease, myocardial infarction, heart failure, and stroke, logistic regression models were fitted within ancestry groups. Where individual-level data were available, case-control sets were constructed using age-balanced designs to mitigate age-related confounding. Results are reported as odds ratios (OR) with corresponding 95% confidence intervals.

For the prediction of cardiometabolic multimorbidity in the UK Biobank Asian subset, gradient boosting models were implemented to evaluate the integrated contribution of GenMetS, age, and socioeconomic and lifestyle factors. Model discrimination was quantified by the area under the receiver operating characteristic curve (AUC), with performance stability assessed across repeated resampling iterations. Model interpretability and the relative importance of individual predictors were evaluated using SHapley Additive exPlanations (SHAP).

In the GUSTO offspring cohort, associations with BMI z-score growth trajectories and MRI-quantified abdominal adiposity were evaluated using standard regression models and group-based comparisons. All statistical modeling and machine learning pipelines were executed using R (v4.1.8) and Python, utilizing the glmnet (v4.1.8), xgboost (v2.0.2), scikit-learn (v1.3.2), and shap (v0.42.1) libraries. Functional annotation and pathway enrichment were conducted through established bioinformatic platforms, as detailed in the **Supplementary Information**.

## Results

### Life-course evaluation of GenMetS across Asian populations

We evaluated GenMetS across cohorts spanning childhood to older adulthood and diverse clinical settings in Singapore, the United Kingdom, Japan, and China, comprising 670,952 participants in total. GenMetS was derived in young adult Singaporean women and evaluated in independent cohorts. We examined whether genetic susceptibility defined in early adulthood is consistently associated with metabolic traits and cardiometabolic outcomes across the life course. Cohort characteristics are summarised in **Tables S1–S3** and **Fig. S1**, and the overall study design is shown in **Figure 1**.

### Genetic ancestry and baseline metabolic burden across cohorts

Genetic ancestry inference identified four major subcontinental groups: Northeast Asian, Southeast Asian, Southwest Asian, and Northern European (**Figure 2a**). These groupings corresponded to self-reported ethnicity and showed clear separation between Asian and European participants. The Singapore discovery cohort was predominantly Northeast Asian (69.2%), with smaller proportions of Southeast Asian (19.5%) and Southwest Asian (11.3%). Similar distributions were observed in the ATTRaCT and GUSTO offspring cohorts. In contrast, the UK Biobank was predominantly Northern European, and its Asian subset was enriched for Southwest Asian ancestry, where this group was approximately five times more prevalent than Northeast Asian. These differences in ancestry composition provide essential context for variation in GenMetS performance across cohorts.

**Fig. 2.**
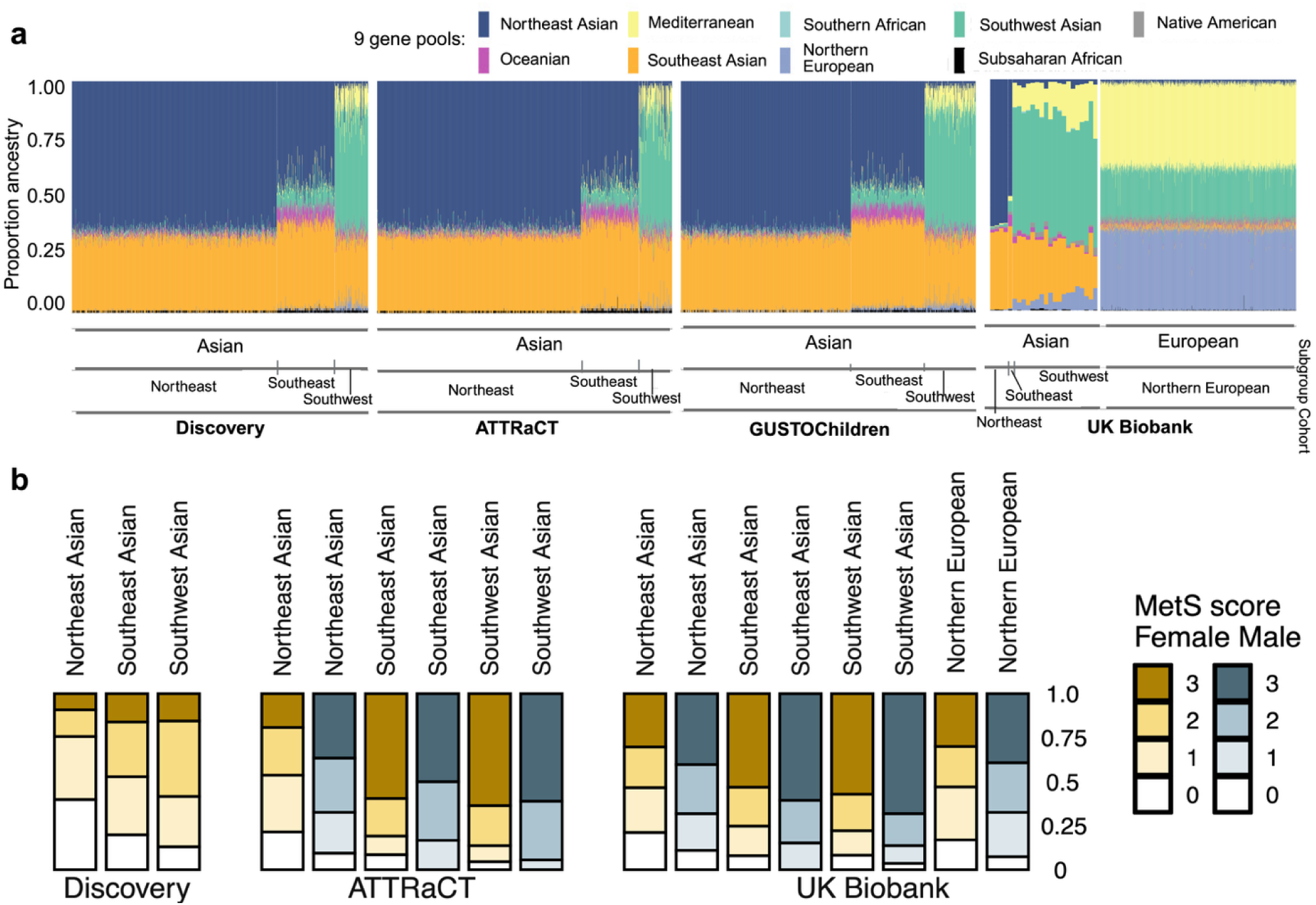
Genetic and metabolic syndrome heterogeneity across study cohorts. **a,** Genetic ancestry of study participants inferred using autosomal ancestry-informative markers to evaluate the representativeness of the GenMetS discovery cohort. Using genetic ancestry-informative markers, individuals were projected onto nine reference ancestral gene pools and classified into four major ancestry groups: Northern European and three Asian groups comprising Northeast Asian, Southeast Asian, and Southwest Asian. All participants (N = 370,246) across the discovery cohorts (S-PRESTO and GUSTO) and validation cohorts (ATTRaCT, GUSTO children, and UK Biobank) were represented by colour-coded admixture proportions based on global ancestral gene pools. For downstream analyses, to maximise statistical power and ensure harmonised cross-cohort comparisons, individuals were analysed in two aggregated ancestry groups: Asian ancestry (all Singapore cohorts and the UK Biobank Asian participants) and European ancestry (the UK Biobank Europeans). **b,** MetS was defined according to the NCEP ATP III criteria and summarised as an ordinal score ranging from 0 to 3, where a score of 3 indicates three or more abnormal components. Stacked bar plots show the proportion of individuals with MetS scores of 0, 1, 2, and 3 across Northeast Asian, Southeast Asian, Southwest Asian, and Northern European ancestry groups in the discovery cohorts (S-PRESTO and GUSTO), the ATTRaCT cohort, and the UK Biobank cohort, stratified by sex.

Baseline metabolic burden also varied across cohorts and age strata (**Figure 2b**). Most women in the Singapore discovery cohorts (S-PRESTO and GUSTO) had low MetS scores, with severe MetS (scores ≥ 3) observed in only 154 participants, consistent with their younger and generally healthy status. In contrast, higher MetS burden was observed in the disease-enriched ATTRaCT cohort while the UK Biobank showed intermediate levels consistent with an older population-based sample. Across cohorts, Southwest Asian participants generally exhibited higher metabolic burden than Northeast Asian participants. Sex differences were most evident in the UK Biobank, where males consistently exhibited higher MetS scores than females. These patterns highlight heterogeneity in cardiometabolic risk across age, ancestry, and clinical context.

### Derivation and biological architecture of GenMetS

GenMetS was constructed using elastic-net regression in a discovery cohort of 1,368 young Singaporean women, leveraging candidate variants derived from 15 polygenic scores across multiple metabolic traits (**Table S6**). The final model retained 4,627 SNPs with non-zero coefficients, reflecting a highly polygenic architecture (**Table S7**). These variants originated from scores spanning glycemic regulation, lipid metabolism, adiposity, and blood pressure (**Fig. S2a**). Overlap of variants across these trait-derived scores indicates that GenMetS reflects shared genetic influences across multiple metabolic pathways rather than a single trait.

To characterize the biological processes represented by the model, variants were mapped to 1,998 genes and evaluated for pathway enrichment. We identified 11 KEGG and 32 Reactome pathways with consistent enrichment across databases (**Fig. S2b**). Enrichment of O-linked glycosylation and voltage-gated calcium channel genes highlights processes previously linked to lipid regulation and cardiovascular risk. As the model is based on autosomal variants underlying core metabolic pathways, cross-sex evaluation is biologically plausible.

### GenMetS explains variance in metabolic traits from childhood to adulthood

We evaluated whether genetic susceptibility defined in early adulthood is consistently associated with MetS and related traits across the life course. Although GenMetS was trained exclusively in women, its construction utilized autosomal variants associated with core metabolic pathways shared between sexes. We therefore evaluated its performance separately in women and men across independent validation cohorts, including the GUSTO child cohort, the ATTRaCT cohort, and the UK Biobank, to assess cross-sex and life-course generalizability (**Figure 3, Table S11**).

**Fig. 3.**
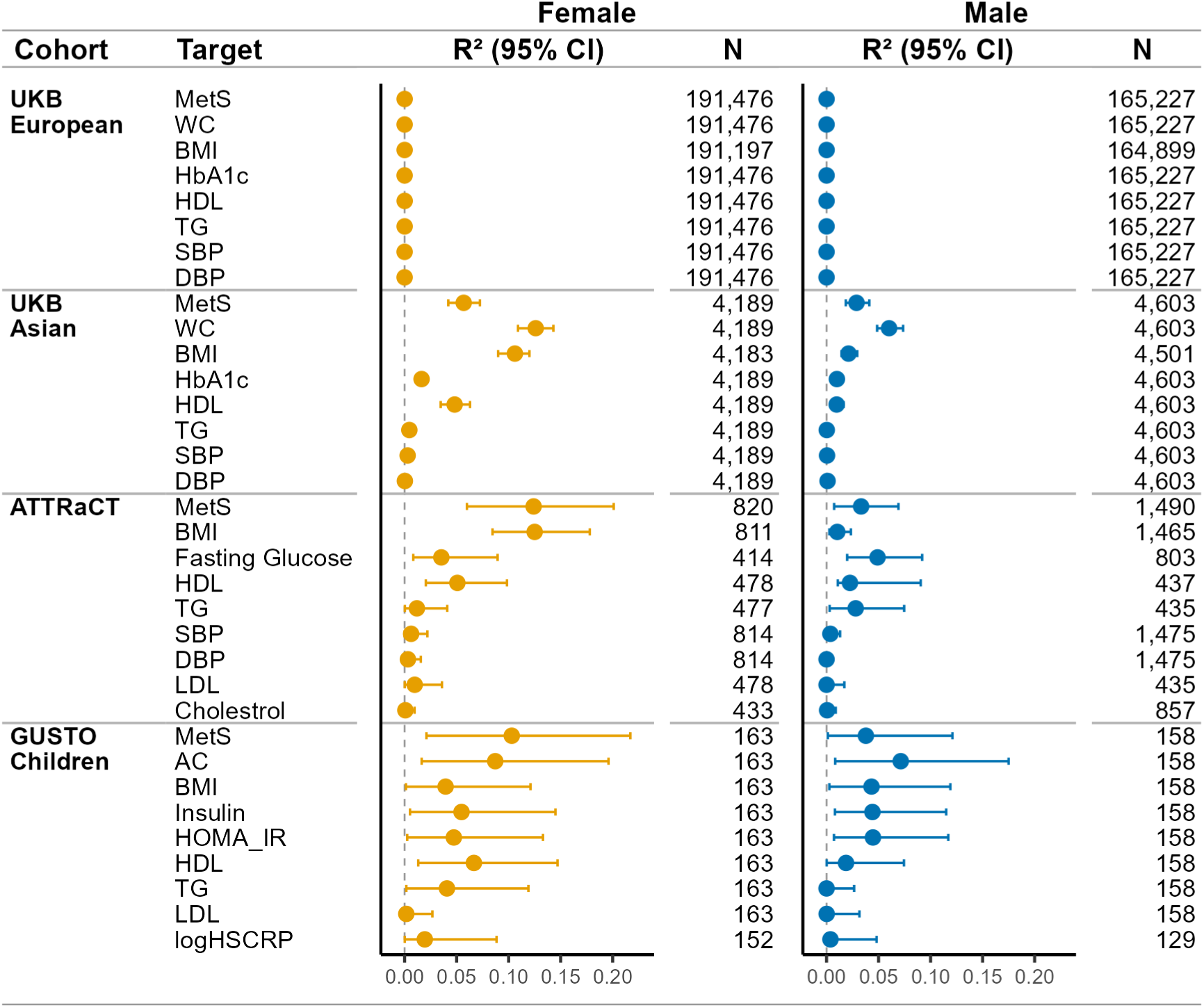
Variance explained by GenMetS for metabolic syndrome and related traits across cohorts. Forest plots display R² values and 95% confidence intervals for the association between GenMetS and clinically measured MetS and related metabolic traits across four cohorts: UK Biobank participants of European ancestry, UK Biobank participants of Asian ancestry, ATTRaCT, and GUSTO children. Results are presented in a sex-stratified manner, with females shown in the left panel and males in the right panel. Female and male categories include women and men in adult cohorts and girls and boys in the child cohort. R² represents the proportion of phenotypic variance explained by GenMetS, with confidence intervals derived from percentile bootstrap resampling (1,000 iterations). Sample sizes for each analysis are provided alongside the corresponding estimates. Metabolic traits evaluated include the composite MetS score, body mass index (BMI), waist circumference (WC), abdominal circumference (AC), systolic blood pressure (SBP), diastolic blood pressure (DBP), triglycerides (TG), fasting glucose, glycated haemoglobin (HbA1c), high-density lipoprotein cholesterol (HDL), low-density lipoprotein cholesterol (LDL), insulin, log10-transformed high-sensitivity C-reactive protein (logHSCRP), and the Homeostatic Model Assessment for Insulin Resistance (HOMA-IR). In GUSTO children, GenMetS was evaluated for abdominal circumference, BMI, and related metabolic traits in both sexes.

Across cohorts of Asian ancestry, GenMetS explained 5.0%–12.4% of the variance in MetS scores among adults and 10.3% in children. The highest variance explained was observed in ATTRaCT women (R² = 0.124) and in girls from the GUSTO cohort (R² = 0.103). Variance explained was lower in UK Biobank Asian women (R² = 0.057), likely reflecting differences in ancestry composition, as the UK Biobank Asian subset is predominantly of Southwest Asian ancestry whereas the discovery cohorts were enriched for Northeast Asian ancestry.

At the trait level, GenMetS explained variance in multiple metabolic components. In UK Biobank participants of Asian ancestry, variance explained for waist circumference was R² = 0.126 in women and 0.060 in men, and in ATTRaCT women, GenMetS explained variance in BMI (R² = 0.125), HDL cholesterol (R² = 0.051), and fasting glucose (R² = 0.035). Across adult cohorts, variance explained was lower in men than in women, whereas in children, sex differences were less pronounced, with GenMetS explaining variance in abdominal circumference in the GUSTO cohort (R² = 0.087 in girls and 0.071 in boys). In UK Biobank participants of European ancestry, GenMetS explained negligible variance in MetS and its component traits (R² < 0.001), suggesting reduced cross-ancestry transferability and consistent with the score capturing genetic architecture more specific to Asian populations.

### GenMetS predicts cardiometabolic disease risk

We next evaluated whether GenMetS was associated with the risk of major cardiometabolic diseases across Asian cohorts. Analyses included T2D, hypertension, coronary artery disease (CAD), myocardial infarction (MI), heart failure (HF), and stroke. Using age-balanced case–control analyses across three cohorts, higher GenMetS was associated with increased odds of all six diseases in both sex-stratified and pooled meta-analyses (**Figure 4**).

**Fig. 4.**
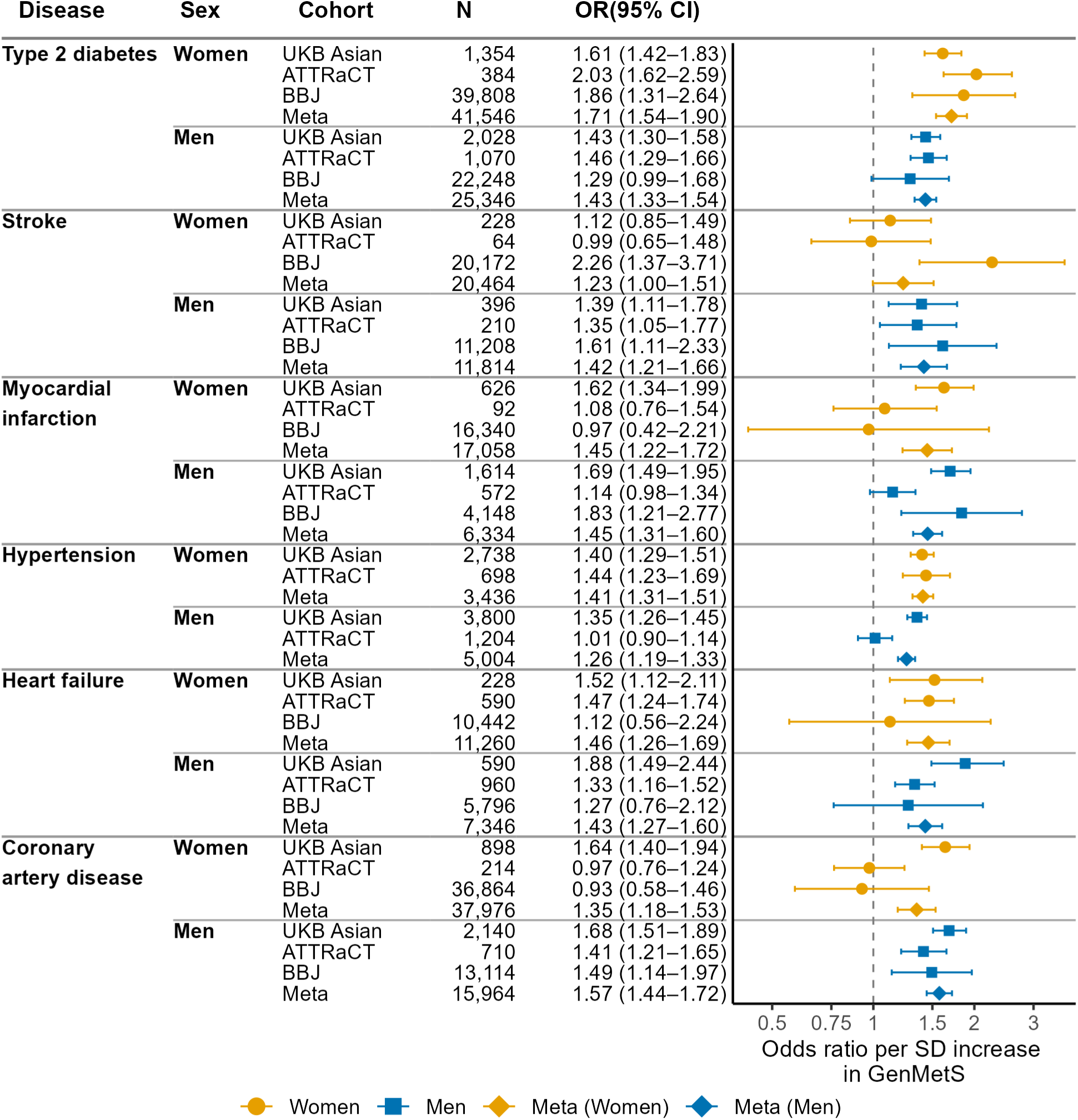
GenMetS prediction of cardiometabolic diseases across three Asian cohorts. Odds ratios (ORs) and 95% confidence intervals (CIs) are shown per one standard deviation (1 SD) increase in GenMetS for six cardiometabolic diseases in participants of Asian ancestry from UK Biobank (UKB Asian), ATTRaCT, and Biobank Japan (BBJ). For each disease, cohort-specific associations are reported separately for women and men, followed by a fixed-effect inverse-variance weighted meta-analysis across cohorts within each sex (Meta, sex-specific). *N* indicates the total number of participants included in each age-balanced case–control analysis, with equal numbers of cases and controls. Points denote cohort-specific estimates and meta-analysed estimates, with horizontal bars indicating 95% CIs. Cohort-specific estimates are shown using sex-specific colours (women, orange; men, blue). Horizontal separators delineate disease groups and sex-specific blocks.

In fixed-effect inverse-variance weighted meta-analyses, a one-standard-deviation (1 SD) increase in GenMetS was associated with higher odds of T2D in both women (OR 1.71, (95% confidence interval (95% CI) 1.54-1.90) ; N = 41,546) and men (1.43 (1.33-1.54); N = 25,346). Associations were also observed for stroke (women, OR 1.23, 95% CI 1.00–1.51; men, OR 1.42, 95% CI 1.21– 1.66), myocardial infarction (women, OR 1.45, 95% CI 1.22–1.72; men, OR 1.45, 95% CI 1.31– 1.60), hypertension (women, OR 1.41, 95% CI 1.31–1.51; men, OR 1.26, 95% CI 1.19–1.33), heart failure (women, OR 1.46, 95% CI 1.26–1.69; men, OR 1.43, 95% CI 1.27–1.60), and CAD (women, OR 1.35, 95% CI 1.18–1.53; men, OR 1.57, 95% CI 1.44–1.72). In CKB, associations were directionally consistent but attenuated (**Fig. S3, Table S12**).

To contextualise these findings, we benchmarked GenMetS against previously published metabolic syndrome polygenic scores [12, 13] and disease-specific polygenic scores [41–45] (**Fig. S4, Table S13**). Benchmarking analyses were performed using the same age-balanced case–control design and fixed-effect meta-analysis across UK Biobank participants of Asian ancestry and the ATTRaCT cohort. Across cardiometabolic outcomes, previously published metabolic syndrome polygenic scores showed modest associations, with odds ratios ranging from approximately 1.01 to 1.34 for Lind2019_MetS [13] and 1.11 to 1.28 for Walree2022_MetS [12] per standard deviation increase, compared with 1.23-1.71 for GenMetS. Disease-specific polygenic scores showed stronger or comparable associations for their corresponding outcomes, including T2D (OR ≈ 1.67-1.74 vs. GenMetS 1.43-1.71), CAD (1.69 vs. GenMetS 1.35-1.57), myocardial infarction (1.32-1.77, vs. GenMetS 1.45), hypertension (1.27-1.35, vs. GenMetS 1.26-1.41) and stroke (1.08-1.43 vs. GenMetS 1.23-1.42) [41–45]. Overall, GenMetS consistently showed stronger associations than conventional metabolic syndrome polygenic scores and demonstrated effect sizes comparable to cardiometabolic disease-specific polygenic scores, despite not being trained on disease endpoints.

### GenMetS provides discrimination for cardiometabolic multimorbidity

We evaluated whether GenMetS predicts cardiometabolic multimorbidity among UK Biobank participants of Asian ancestry. The analytic sample included 1,045 cases (336 women and 709 men) and 3,926 controls (1,937 women and 1,989 men) with complete diagnostic and covariate data. Sex-stratified XGBoost models were trained using three predictor configurations: (Model 1) age and GenMetS; (Model 2) age and socioeconomic and lifestyle factors; and (Model 3) an integrated model including all predictors (**Figure 5a–d**).

**Fig. 5.**
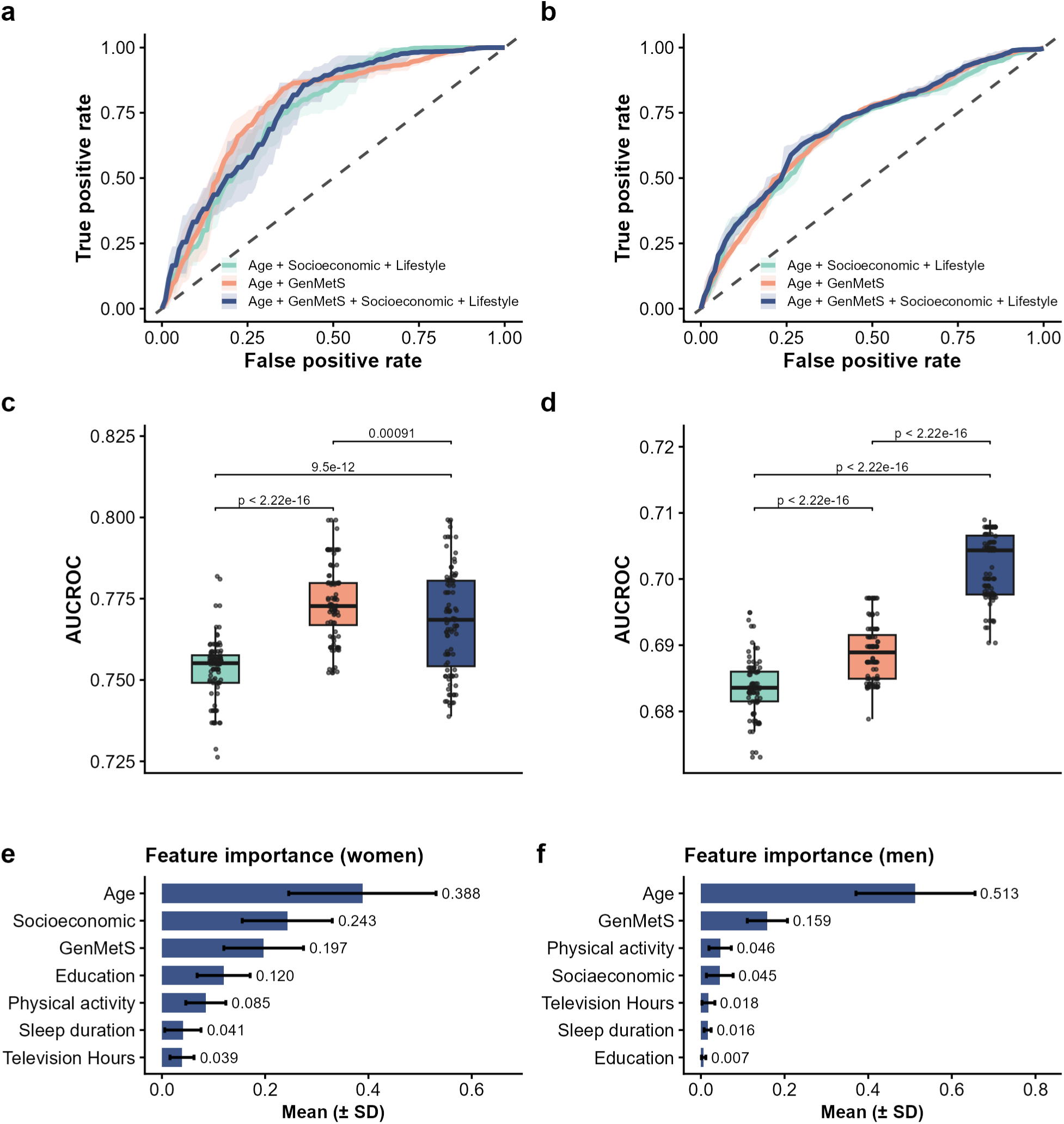
Prediction of cardiometabolic multimorbidity using GenMetS, socioeconomic and lifestyle factors. Receiver operating characteristic (ROC) analyses were performed using XGBoost models in UK Biobank participants of Asian ancestry (N = 4,971; 1,045 cases and 3,926 controls). Three predictor configurations were evaluated: age with socioeconomic and lifestyle factors (education, socioeconomic deprivation measured by the Townsend Deprivation Index, physical activity, television viewing time, and sleep duration); age with GenMetS; and age with GenMetS together with socioeconomic and lifestyle factors. The dashed diagonal line indicates no discrimination (area under the curve (AUC) = 0.5). Model robustness was assessed using 100 bootstrap iterations, and mean AUC values with standard deviations are reported. Relative predictor contributions were quantified using SHAP, with mean absolute SHAP values averaged across bootstrap iterations. **a,** ROC curves showing prediction performance of the three models in women, with AUC values of 0.753 for age with socioeconomic and lifestyle factors, 0.774 for age with GenMetS, and 0.768 for the integrated model. **b,** ROC curves showing prediction performance of the three models in men, with AUC values of 0.683, 0.689, and 0.702 for the same three models, respectively. **c,** Distribution of bootstrap AUC values across 100 iterations for women, illustrating model variability and pairwise model comparisons using paired t-tests. **d,** Distribution of bootstrap AUC values across 100 iterations for men, illustrating model variability and pairwise model comparisons using paired t tests. **e,** Relative predictor contributions in women derived from SHAP analysis of the integrated risk model. **f,** Relative predictor contributions in men derived from SHAP analysis of the integrated risk model.

In women, the model including age and GenMetS showed higher discrimination than the model including age and socioeconomic and lifestyle factors (mean (SD) AUC 0.774 (0.012) vs 0.753 (0.009), paired t-test across bootstrap iterations, P < 2.2 × 10⁻¹⁶). Adding socioeconomic and lifestyle factors to age and GenMetS did not increase AUC (0.774 (0.012) vs 0.768 (0.016); **Figure 5a, c**). In men, the model including age and GenMetS showed slightly higher discrimination than the model including age with socioeconomic and lifestyle factors (AUC 0.689 (0.004) vs 0.683 (0.004), P < 2.2 × 10⁻¹⁶). The integrated model showed higher discrimination than the model including age with socioeconomic and lifestyle factors (AUC 0.702 (0.005) vs 0.683 (0.004); **Figure 5b, d**).

SHAP analysis of the integrated model ranked age as the top contributor, followed by GenMetS. Mean (SD) SHAP values were 0.388 (0.143) for age and 0.197 (0.077) for GenMetS in women, and 0.513 (0.142) and 0.159 (0.048), respectively, in men. Socioeconomic and lifestyle variables showed smaller contributions **(Figure 5e, f).**

### Childhood GenMetS predicts obesogenic growth patterns and abdominal adiposity

We evaluated whether GenMetS calculated from children’s genotypes was associated with early-life growth patterns and abdominal adiposity in the GUSTO children cohort. Five previously defined BMI z-score trajectories from birth to age six in GUSTO children (N = 994) were examined [46]. These trajectories included two obesogenic patterns including early acceleration, characterised by rapid BMI gain from birth, and late acceleration, where BMI begins to accelerate after one year of age. The remaining three trajectories were stable normal high, stable normal, and stable normal low, representing normal, non-obesogenic growth patterns. Together, these five trajectories capture the full range of early childhood BMI development from 0 to 6 years in this cohort (**Figure. 6a**). GenMetS differed significantly across childhood BMI trajectory groups (ANOVA P = 1.2 × 10⁻⁹; **Figure. 6b**) and was significantly higher in children exhibiting early-acceleration and late-acceleration growth patterns compared with those following the stable normal trajectory (*P* = 5.7 × 10⁻^7^ and 4.3 × 10⁻⁵, respectively, **Figure. 6b**). In a subset of children who underwent abdominal magnetic resonance imaging at age six, higher GenMetS values were associated with greater abdominal adiposity, including increased superficial subcutaneous fat, deep subcutaneous fat, and intra-abdominal fat compartments (**Figure. 6c, d**).

**Fig. 6.**
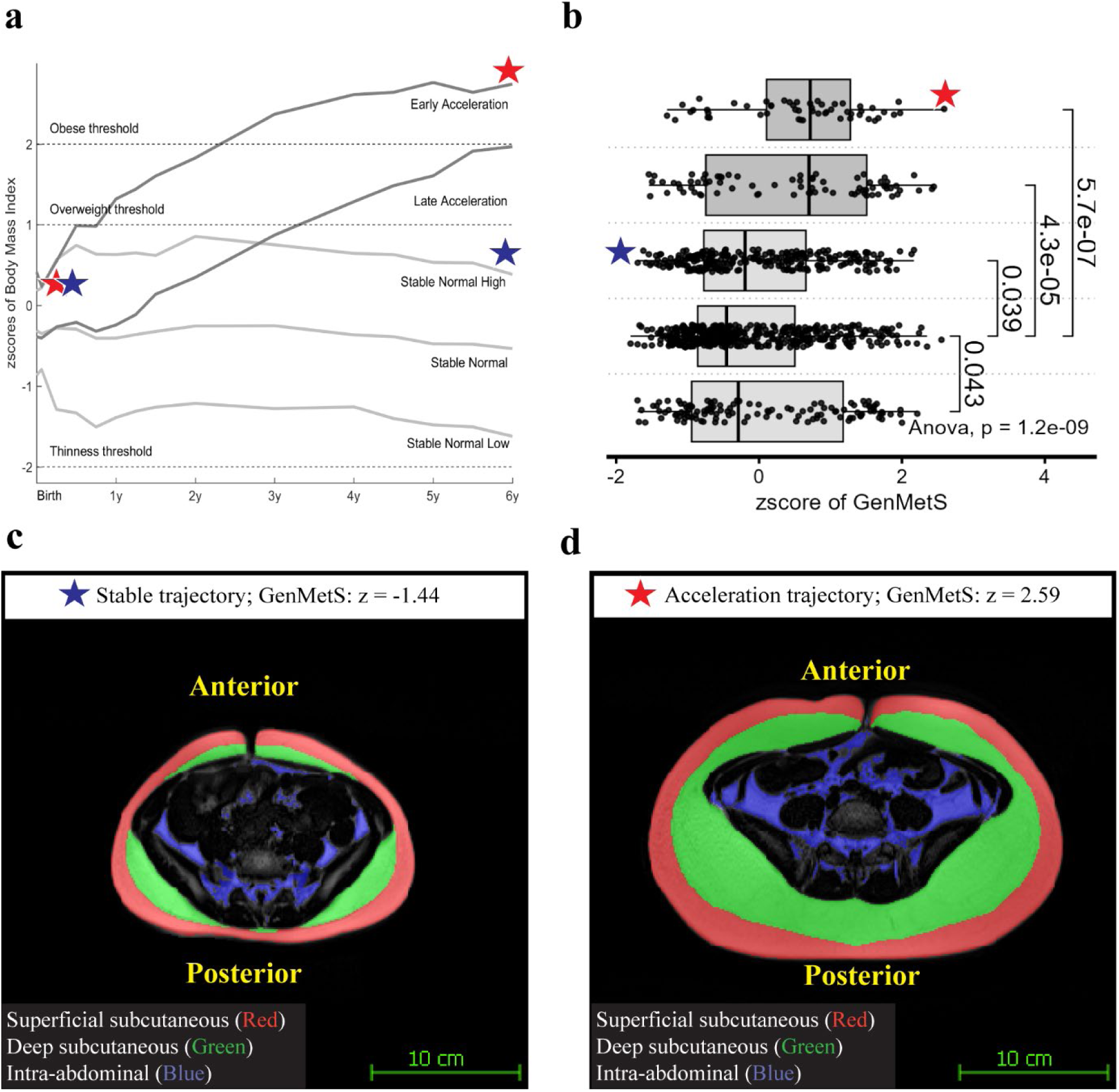
Higher GenMetS is associated with obesogenic growth patterns and abdominal adiposity in children. **a,** Five distinct growth trajectories derived from BMI z-scores measured between birth and age 6 years in the GUSTO cohort. Two trajectories reflect obesogenic growth patterns. The early acceleration trajectory is characterised by rapid BMI gain from birth, whereas the late acceleration trajectory shows BMI beginning to accelerate after approximately one year of age. The remaining three trajectories, stable normal high, stable normal, and stable normal low, represent non-obesogenic growth patterns. An illustrative example highlights two children marked by blue and red stars with similar birth weights (blue, 2.94 kg, z = −0.02; red, 3.18 kg, z = 0.39) who diverged by age 6 years. The child indicated by the blue star remained on the normal trajectory, whereas the child marked by the red star transitioned to early acceleration trajectory. **b,** GenMetS calculated from child genotypes differed significantly across the five growth trajectories (ANOVA P = 1.2 × 10^-9^). Each boxplot of GenMetS values corresponds to each BMI trajectory. Children following the early acceleration and late acceleration trajectories exhibited higher GenMetS compared with those following the stable normal trajectory (P = 5.7 × 10^-7^ and 4.3 × 10^-5^, respectively). The blue and red stars denote the corresponding children highlighted in panel a, representing individuals with high (2.59) and low (-1.44) GenMetS z-scores. **c-d,** Representative axial abdominal magnetic resonance images obtained at age 6 years illustrate differences in abdominal adiposity between the highlighted children. Images are presented at identical magnification to enable direct comparison. Sample sizes are indicated in the figure.

## Discussion

In this study, we show that the life stage at which genetic liability is modelled influences the biological signal captured by polygenic scores. By deriving GenMetS in early adulthood, a period of relative physiological stability with reduced clinical confounding, we identified a genetic signal that remains consistently associated with metabolic traits, cardiometabolic diseases, and multimorbidity across the life course in Asian populations. GenMetS explained 5.0–12.4% of the variance in metabolic syndrome in adults and 10.3% in children and showed consistent associations with six cardiometabolic diseases. Notably, these associations were detectable in early childhood, where GenMetS was linked to obesogenic growth trajectories and abdominal adiposity, and persisted into older adulthood. Performance was attenuated in European populations, consistent with ancestry-specific genetic architecture. Despite being derived in women, associations were consistent across sexes. Together, these findings indicate that genetic liability modelled in early adulthood captures a stable biological signal that is detectable from early life and remains relevant across the life course.

A key feature of GenMetS is its derivation from metabolic phenotypes in early adulthood. Most existing polygenic scores for cardiometabolic traits are derived in middle-aged or older populations, where genetic effects are influenced by medication use, chronic disease, and long-term environmental exposures. In contrast, modelling genetic liability in early adulthood captures genetic effects under conditions of reduced clinical confounding, reflecting upstream metabolic regulation rather than downstream disease processes. The consistent associations observed from childhood to older adulthood indicate that this early-defined genetic signal remains stable across the life course. Although direct comparisons with scores derived in later adulthood were not performed, our findings support a temporal dimension in polygenic modelling, whereby the life stage of model derivation influences the biological signal captured. Together, these results indicate that genetic liability to metabolic dysfunction is shaped by both ancestry and the timing of model construction.

GenMetS showed marked ancestry-related differences in predictive performance, with strong performance in Asian populations but limited transferability to European populations. This pattern is consistent with established effects of genetic distance, allele frequency differences, and linkage disequilibrium structure on polygenic score portability. Although the model incorporates variants identified across multiple ancestries, SNP weights were estimated in Asian cohorts, resulting in a genetic signal that is optimized for Asian-specific architecture. Notably, performance also varied within Asian populations, reflecting subcontinental heterogeneity. The attenuated performance observed in the UK Biobank Asian subset, which includes a higher proportion of Southwest Asian ancestry compared with the Northeast and Southeast Asian composition of the Singapore cohorts, highlights the importance of fine-scale ancestry in polygenic modelling. These findings indicate that both cross-continental and within-Asia genetic heterogeneity influence model performance and should be explicitly considered in the development and application of polygenic scores.

A further strength of GenMetS lies in its multivariable design. Because cardiometabolic diseases arise from interacting physiological traits, composite models may better reflect underlying biological architecture than single-trait scores [47]. GenMetS was constructed by integrating information from 15 polygenic scores representing individual metabolic traits, thereby capturing shared genetic liability across these domains. This design may help explain why GenMetS showed stronger associations with cardiometabolic outcomes than conventional metabolic syndrome polygenic scores and comparable associations to several disease-specific scores (**Fig. S5 and S6**).

In multimorbidity analyses, models combining age and GenMetS achieved similar or higher discrimination than models incorporating age with demographic, socioeconomic, and lifestyle factors, while the addition of these variables to age and GenMetS provided minimal incremental benefit. These results indicate that GenMetS captures risk components not fully reflected by conventional covariates. SHAP analysis further supported this interpretation, ranking age as the primary contributor, followed by GenMetS, with substantially smaller contributions from socioeconomic and lifestyle factors. Together, these findings support the role of genetically informed measures as complementary predictors in cardiometabolic risk stratification and suggest that models derived under minimal confounding may enable earlier identification of individuals at risk, supporting life-course–oriented prevention strategies.

Pathway enrichment analyses showed that variants contributing to GenMetS are enriched in biological processes related to extracellular matrix organisation, O-linked glycosylation, and calcium signalling [48–51], which have established roles in lipid metabolism, vascular function, and metabolic regulation. Genes mapped to GenMetS variants included both established cardiometabolic loci (CETP, FTO, GCK, LPL) and less-characterized genes such as MED23 and MYO1F, indicating that the model captures both known and potentially underexplored components of metabolic biology [14, 52].

Several limitations should be considered. The discovery cohort was modest in size and restricted to women, which may limit generalizability and contribute to lower performance in men and other ancestry groups. Although evaluated in large external cohorts with sex-stratified analyses, further validation in more diverse populations is needed. Performance differences across Asian cohorts reflect sub-population heterogeneity and highlight the need for broader regional genomic resources. In addition, harmonisation of metabolic phenotypes across cohorts required the use of proxies, such as BMI for waist circumference, which may introduce measurement error. Finally, prospective longitudinal studies are needed to evaluate the clinical utility of GenMetS beyond existing risk assessment approaches.

### Conclusions

GenMetS is a polygenic measure of metabolic liability derived in early adulthood that captures a stable genetic signal associated with metabolic traits and cardiometabolic outcomes across the life course in Asian populations. This signal is detectable from early childhood and persists into adulthood across multiple disease outcomes. These findings demonstrate that the life stage of model derivation, alongside ancestry, shapes the biological signal captured by polygenic scores, and support the development of life-stage- and ancestry-informed approaches for cardiometabolic risk assessment and prevention. This framework supports the development of next-generation polygenic models that integrate temporal and ancestry dimensions to improve risk prediction and prevention.

## List of abbreviations

MetS: Metabolic syndrome
SNP: Single nucleotide polymorphism
HDL(-C): high-density lipoprotein cholesterol
DBP: diastolic blood pressure
SBP: systolic blood pressure
T2D: Type 2 Diabetes
CAD: coronary artery disease
NAFLD: Non-alcoholic fatty liver disease
OR: odds ratio.

## Supplementary Material

Please refer to the separated document of **Supplementary Information.**

## Ethics approvals

Ethical approval was obtained from the SingHealth Centralized Institutional Review Board (reference numbers 2019/2655E and 2019/2143D), and written informed consent was obtained from all participants in the GUSTO and S-PRESTO cohorts. The S-PRESTO study is registered at ClinicalTrials.gov (NCT03531658).

This study used data from the UK Biobank (application number 85457), the China Kadoorie Biobank, the Biobank Japan Project, and the ATTRaCT cohort under approved data access. The UK Biobank study was approved by the North West Multi-centre Research Ethics Committee (REC reference: 11/NW/0382); the China Kadoorie Biobank by the Oxford Tropical Research Ethics Committee and the Chinese Center for Disease Control and Prevention Ethical Review Committee; and the Biobank Japan Project by the ethics committees of the Institute of Medical Science, The University of Tokyo, and participating institutions. All participants provided written informed consent.

## Data availability

MetS related SNPs were compiled by consolidating SNP identifiers from 15 polygenic scores meticulously selected from the Polygenic Score (PGS) Catalogs (https://www.PGSCatalog.org, with the index information including PGS000828, PGS001227, PGS000661, PGS000891, PGS000686, PGS000659, PGS000699, PGS000306, PGS000684, PGS000685, PGS001133, PGS000302, PGS000301, and PGS001134, **Table S6**). For cohort-specific datasets, each individual cohort has to be contacted at different data access policies. Individual-level GUSTO and S-PRESTO data are available under restricted access administered by GUSTO vault (https://gustodatavault.sg/). Access can be obtained by application at https://fas.sicsapps.com/site/login. Individual-level data from ATTRaCT cohort can be accessed by contacting the steering committee through the authors. Access to UK Biobank data can be obtained by application to UK Biobank (https://www.ukbiobank.ac.uk/).

## Authors’ contributions

Conceptualization: H.P., D.W. Supervision: D.W. Formal analysis: H.P., D.W., E.L., Y.C., S.S.E.L., A.L.T., Kazuo M., E.E., K.L., and R.G.W. Resources: S.S.E.L, R.F., S.A.S., S.S.V., K.I., Koichi M., K.M.L.T., K.H.T., J.K.C., C.L., A.M.R, K.L., R.G.W., and J.G.E. Investigation: D.W., H.P, E.L., V.G., S.S.E.L., R.F., J.G.E., K.H.T., J.K.Y.C., D.C.S., Koichi M. and K.I. Visualization: H.P., E.L., A.L.T., P.F.T., X.Z. and Y.C. Funding acquisition: H.P., D.W. Early ideation/conceptualization: K.M.L.T. and D.C.S. Writing of original draft: H.P., D.W., E.L., J.H., Y.C., A.L.T, S.S.E.L., R.F., V.G, Kazuo M. K.I, K.L. and R.G.W. Writing, review and editing: all authors revised the manuscript critically for important intellectual content; and gave final approval of the version submitted for publication; and agreed to be accountable for all aspects of the work in ensuring that questions related to the accuracy or integrity of any part of the work were appropriately investigated and resolved.

## Acknowledgements

This work was supported by the A*STAR Computational Resource Centre through the use of its high-performance computing facilities. Acknowledgements for each study team and ethical approval information can be found in **Supplementary Information**.

