## Supplementary Methods and Figures for "Genetic liability to metabolic dysfunction modelled in early adulthood predicts cardiometabolic risk across the life course in Asian populations"

### Discovery cohort

#### S-PRESTO

S-PRESTO is a Singapore preconception cohort of young adult women [1]. For this study, age, self-reported ethnicity, and metabolic trait measurements from the initial preconception visit were used. Genotype data were generated from peripheral blood DNA using whole-genome sequencing on the Illumina HiSeq X platform. A total of 934 women had complete genotype and metabolic trait data and were included.

#### GUSTO mothers

GUSTO is a longitudinal mother-offspring cohort established in Singapore [2]. For this study, maternal demographic variables and metabolic trait measurements collected at the 8-year postpartum follow-up were used. Maternal DNA was extracted from blood collected at mid-gestation and genotyped using the Illumina OmniExpress plus Exome array, followed by imputation using the 1000 Genomes Phase 3 reference panel [3] . Complete genotype and metabolic trait data were available for 434 mothers.

#### Genotype quality control for the discovery cohorts

Genotype data from unrelated women in S-PRESTO (N = 934) and GUSTO mothers (N = 434) were merged using shared variants. Variants were retained after applying the following criteria: SNP missingness < 5%, minor allele frequency > 0.05, Hardy–Weinberg equilibrium (HWE) P-value > $1 \times 10^{-6}$, and exclusion of variants within the major histocompatibility complex region (hg19 chr6:25477797-36448354). This yielded 4,946,168 biallelic variants.

Genotype data were generated using cohort-specific platforms. In S-PRESTO, peripheral blood DNA underwent whole-genome sequencing on the Illumina HiSeq X platform. In GUSTO, genotype data were obtained from blood samples collected at mid-gestation using the Illumina OmniExpress plus Exome array, followed by imputation with the 1000 Genomes Phase 3 reference panel [3, 4]. Standardised genotype quality control procedures were applied across cohorts. Single-nucleotide polymorphisms (SNPs) were retained if they met the following criteria: (1) SNP missingness < 5%, (2) minor allele frequency > 0.05, and (3) Hardy-Weinberg equilibrium P-value > $1\times 10^{-6}$, exclusion of variants within the major histocompatibility complex region (hg19 chr6:25477797-36448354). This yielded 4,946,168 biallelic variants in GRCh37 reference genome build.

### Validation cohorts

#### GUSTO children

Children from the GUSTO cohort were included as an independent validation cohort. Anthropometric measurements were obtained longitudinally from birth to six years and fasting biochemical and blood pressure measures were collected at the 6-year follow-up visit [5-7]. Child DNA was obtained from cord tissue or peripheral blood [8] and genotyped using the Illumina OmniExpress plus Exome array, followed by imputation using the 1000 Genomes Phase 3 reference panel. Complete genotype and metabolic trait data were available for 500 children.

At age 6 years, abdominal circumference (AC) was measured [6] and fasting venous blood samples were collected to assess fasting glucose (FG), serum insulin, total cholesterol (TC), high-density lipoprotein cholesterol (HDL) and triglycerides (TG). Homeostasis model assessment of insulin resistance (HOMA-IR) was calculated using the formula [9]: [fasting insulin (mU/L) * fasting glucose (mmol/L)] / 22.5. Systolic blood pressure (SBP) and diastolic blood pressure (DBP) were measured from the right upper arm by trained research coordinators [5] . Paediatric metabolic syndrome scores were calculated using the formula introduced in [10], as zAC + (zSBP + zDBP)/2 + (zTG + zHDL)/2 + zHOMA-IR, where z-scores of the corresponding variables were used.

Genotype quality control for child data followed criteria similar to those used in the discovery cohorts. SNPs were excluded if the minor allele frequency (MAF) was below 5%, the call rate was below 95%, or the HWE P-value was less than $1\times10^{-6}$. The resulting genotype dataset was mapped to the GRCh37 genome assembly.

#### ATTRaCT

The Asian neTwork for Translational Research and Cardiovascular Trials (ATTRaCT) is a Singapore-based cohort of Asian adults aged 22–94 years and enriched for heart failure cases [11, 12]. The study included deep phenotyping and whole-genome sequencing in 2,445 participants. Whole-genome sequencing was performed on the Illumina HiSeq 4000 and HiSeq X platforms, with reads aligned to the GRCh38 reference genome. Because waist circumference was unavailable, the adiposity component of MetS was defined using BMI > 25 kg/m$^2$ [14,15]. In 81 participants with both BMI and waist circumference data, this proxy showed approximately 70% concordance for obesity classification in MetS assessment.

Quality control on ATTRaCT geneotype data was performed using PLINK 1.9. Samples were excluded for call rate < 95%, sex discordance, or high heterozygosity. SNPs were excluded for missingness > 5%, HWE P-value < $1\times10^{-6}$, or minor allele frequency < 0.01.

#### UK Biobank (UKB)

UK Biobank is a large population-based cohort comprising 502,408 participants [13]. For this study (application ID 85457), records were updated in September 2023 after accounting for participant withdrawals. Imputed genotype data from release version 3 were used, and analyses were restricted to unrelated participants with complete data on metabolic traits, age, sex, and ethnic background who passed sample quality control. Based on self-reported ethnic background (field 21000), the final analytic sample included 8,792 participants of Asian ancestry and 356,703 self-reported British participants (total N = 365,495; **Fig. S1**).

Disease information was derived from International Classification of Diseases, Tenth Revision (ICD-10) codes recorded in linked electronic health records in the UK Biobank, retrieved in August 2024. Disease cases were extracted from UK Biobank field 41270 (ICD-10 diagnoses). Disease definitions are detailed in **Table S9**.

Type 2 diabetes (T2D) cases included participants with ICD-10 code E11 (non-insulin-dependent diabetes mellitus), while controls excluded codes E10, E12, E13, and E14. Hypertension (HTN) cases included participants with ICD-10 codes I10 (essential hypertension) and I15 (secondary hypertension). Heart failure (HF) cases included participants with ICD-10 codes I11.0, I13.0, I13.2, I25.5, I42.0, I42.5, I42.8, I42.9, I50.0, I50.1, and I50.9, based on the latest report [14]. Non-alcoholic fatty liver disease (NAFLD) cases included participants with ICD-10 codes K76.0 and K75.8, while excluding those with alcoholic liver disease, viral hepatitis, autoimmune liver disease, and other competing liver conditions defined by ICD-10 codes K70, B16, B17, B18, B19, K83.0A, K83.0F, K74.3, K75.4, E83.1, E83.0B, E88.0A, E88.0B, I82.0, K76.5, K73.9, K73.2, K74.4, and K74.5 [15]. Myocardial infarction (MI) cases included participants with ICD-10 codes I20. Stroke cases were identified using ICD-10 codes I60, I61, I63, I64, H34.1, and G45 [16]. Coronary artery disease (CAD) was defined as myocardial infarction, chronic ischaemic heart disease, or angina using ICD-10 codes I20-I25, based on the latest CAD GWAS study [17].

Six sociodemographic and lifestyle variables were included in multimorbidity analyses: age, education, socioeconomic status, television viewing time, physical activity, and sleep duration. Physical activity was assessed using adapted questions from the short International Physical Activity Questionnaire, covering the frequency, intensity, and duration of walking and moderate and vigorous activity. Activity was converted into total physical activity expressed as metabolic equivalent task minutes per week (MET-min/week; field 22040) [18]. Television viewing was assessed using the question, “In a typical day, how many hours do you spend watching television?” (field 1070). Sleep duration was assessed using the question, “About how many hours of sleep do you get in every 24 h?” (field 1160). The Townsend Deprivation Index (field 22189) was used as a measure of socioeconomic deprivation, with higher values indicating greater deprivation. Educational attainment was derived from qualification data (field 6138). Statistical details for these variables in the multimorbidity subset are reported in **Table S9**.

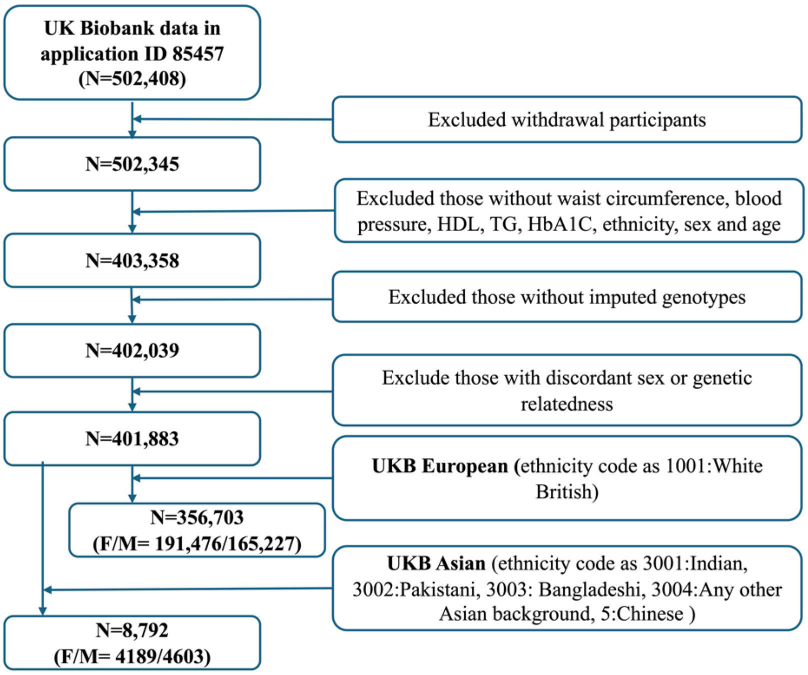


**Fig. S1: Inclusion criteria for UK Biobank.** F: female; M: male; N: number.

### Biobank Japan (BBJ)

Biobank Japan (BBJ) is a hospital-based national biobank project that collects DNA, serum samples, and clinical information from 12 medical institutes in Japan [19, 20]. Approximately 200,000 patients were enrolled, and all participants provided written informed consent approved by the ethics committees of the Institute of Medical Sciences, the University of Tokyo, and the RIKEN Center for Integrative Medical Sciences.

Participants were genotyped using the Illumina HumanOmniExpress Genotyping BeadChip or a combination of the Illumina HumanOmniExpress and HumanExome BeadChips. Sample quality control excluded closely related individuals based on identity-by-descent analysis, as well as individuals with a sample call rate < 0.98, heterozygosity rate > 4 standard deviations, or principal component outlier status relative to East Asian clusters. Variant quality control excluded SNPs with call rate < 0.99, minor allele frequency < 0.01, or HWE P < 1.0 × 10^-6. Genotypes were pre-phased using EAGLE and imputed using minimac3 with the 1000 Genomes Project Phase 3 reference panel together with an in-house Japanese whole-genome sequencing reference panel from BBJ (N = 1,037).

### China Kadoorie Biobank (CKB)

The China Kadoorie Biobank (CKB) is a prospective cohort comprising 512,724 adults aged 30-79 years recruited between 2004 and 2008 from ten geographically diverse regions across China, including five urban and five rural areas. Details of the study design and data collection have been described previously [21]. For the present study, genotype data and adjudicated disease outcomes were used for evaluation of GenMetS. Disease definitions were based on ICD-coded outcomes together with adjudicated clinical endpoints, as defined by the CKB study.

**MetS definition**

MetS was defined according to the National Cholesterol Education Program Adult Treatment Panel III (NCEP ATP III) criteria [22] as the presence of three or more of the following components: (i) elevated waist circumference (≥80 cm in Asian women or ≥90 cm in Asian men; ≥88 cm in non-Asian women or ≥102 cm in non-Asian men); (ii) triglycerides ≥1.7 mmol/L or lipid-lowering treatment; (iii) HDL cholesterol <1.29 mmol/L in women or <1.03 mmol/L in men, or lipid-lowering treatment; (iv) fasting glucose ≥5.6 mmol/L or glucose-lowering treatment; and (v) systolic blood pressure ≥130 mmHg or diastolic blood pressure ≥85 mmHg, or antihypertensive treatment. Each abnormal component was coded as 1 and summed to derive a MetS score ranging from 0 to 5. Because fewer than 5% of participants exhibited four or more abnormal components in discovery cohort, MetS scores ≥3 were truncated to 3 to stabilise variance while preserving the ordinal ranking of metabolic burden. MetS scores were used as the outcome for genetic modelling and are summarised across cohorts and age groups in **Table S5**.

**The GenMetS SNP distribution from the GWAS resources**

Among the 4,627 SNPs retained in GenMetS, each variant was traced back to its source GWAS-derived polygenic score and summarised using an UpSet plot (**Fig. S2a**). Source scores covered waist circumference (WC) (PGS000828 [23] and PGS001227 [24]), low-density lipoprotein cholesterol (LDL) (PGS000661 [25] and PGS000891 [26] ), high-density lipoprotein cholesterol (HDL) (PGS000660 [27] and PGS000686 [28]), triglycerides (PGS000659 [25] and PGS000699[28]), fasting glucose (PGS000306 [29] and PGS000684 [28]) and diastolic blood pressure (DBP) (PGS001133 [24] and PGS000302 [29]) and systolic blood pressure (SBP) (PGS000301 [29], PGS001134 [24]), **Table S6.** Variants contributing to GenMetS originated from all component traits, with substantial sharing across trait-derived scores, indicating that GenMetS captures genetic influences across multiple metabolic domains rather than being driven predominantly by a single trait.

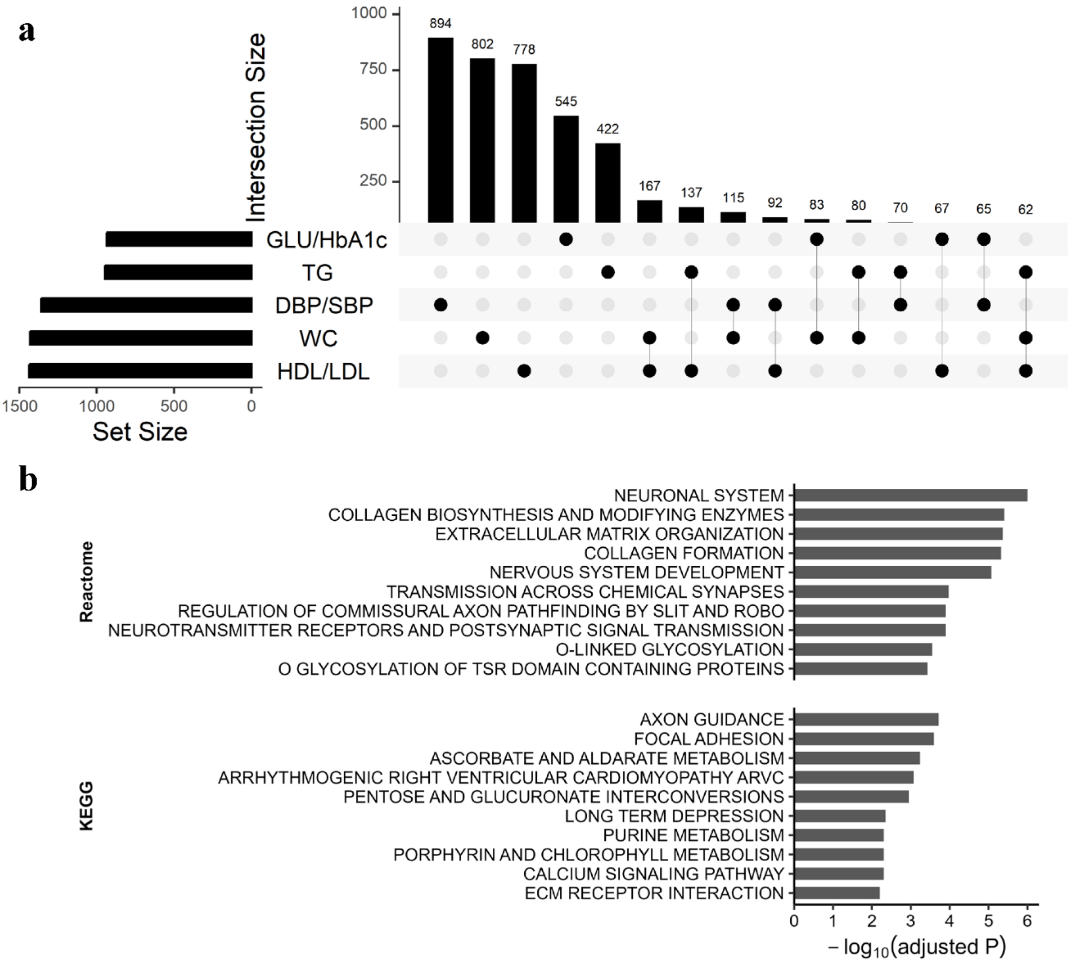


**Fig. S2, Genetic architecture and functional enrichment of variants included in GenMetS. a,** UpSet plot illustrating the compositional origin of the genetic variants included in GenMetS. The selected SNPs were derived from polygenic risk scores (PRS) for all five metabolic traits (GLU/HbA1c, TG, DBP/SBP, WC, and HDL/LDL). Vertical bars indicate the number of SNPs shared across trait-derived PRSs, whereas horizontal bars represent the total number of SNPs contributed by each trait. Connected dots denote the specific traits from which the variants were selected, demonstrating the multi-trait genetic architecture captured by GenMetS. **b,** Top enriched pathways by FUMA. Pathway enrichment analysis of genes mapped from GenMetS variants based on KEGG and Reactome databases. Bar lengths correspond to enrichment significance expressed as $-\log_{10}$(adjusted *P*) after FDR correction. Values were capped at a predefined threshold to facilitate visualization. Redundant pathways were consolidated through clustering, and representative pathways are displayed. The enriched pathways highlight shared biological mechanisms underlying metabolic dysregulation.

**Pathway analysis of SNPs in GenMetS**

The 4,627 SNPs retained in GenMetS were annotated using the biomaRt package [30]. Of these, 2,304 SNPs mapped to Ensembl genes, corresponding to 1,998 unique genes. These genes were analysed using the GENE2FUNC module in FUMA [31] for functional annotation and pathway enrichment. Gene-set enrichment analysis was performed using

KEGG and Reactome databases. We required at least five overlapping genes per gene set and an adjusted P-value below 0.01 for significance. 11 KEGG pathways and 32 Reactome pathways were significantly enriched and the top 10 of KEGG and Reactome pathways were shown in **Fig. S2b**. Enriched pathways involved neuronal signalling, extracellular matrix organisation, calcium signalling, and metabolic processes, indicating that GenMetS captures genetic signals across multiple biological domains relevant to cardiometabolic regulation.

**GenMetS predicts cardiometabolic diseases in CKB**

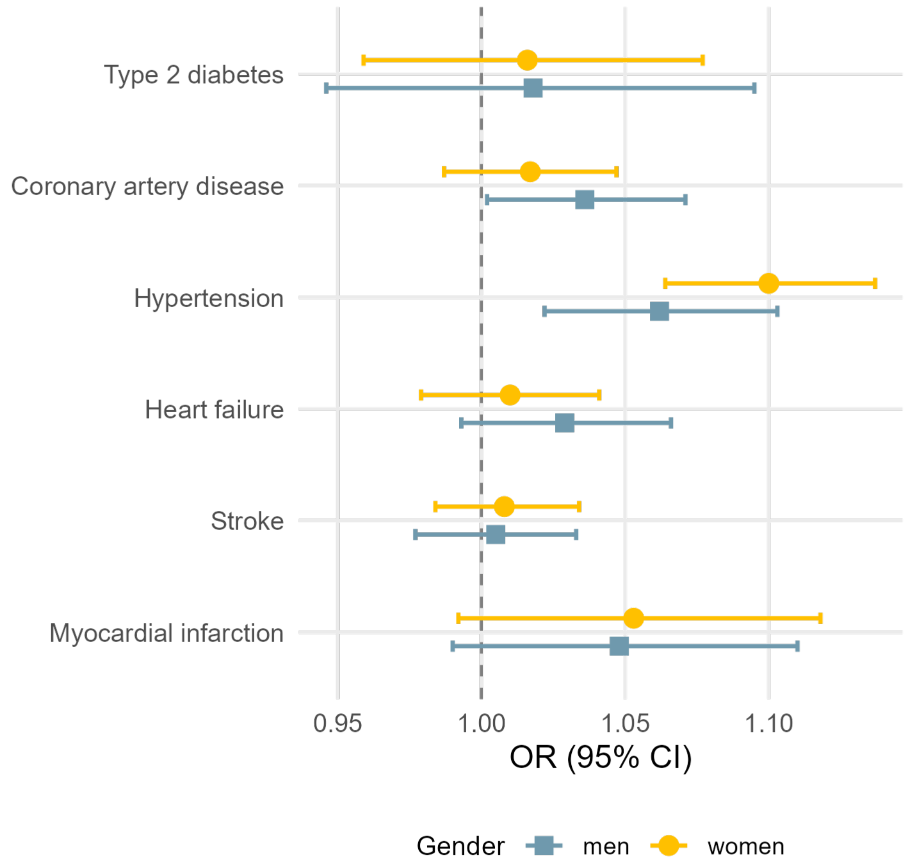


**Fig. S3:** GenMetS in the prediction of six cardiometabolic diseases across CKB in different sex group. Squares and circles show the odds ratios (OR) for women and men, and horizontal bars show the 95% confidence intervals (95% CI). Odds ratios are per standard deviation increase of the estimated GenMetS scores. The detailed results are in **Table S12**.

**Benchmarking GenMetS against metabolic syndrome and disease-specific polygenic scores**

To benchmark the disease relevance of GenMetS, we compared its associations with cardiometabolic outcomes against previously published metabolic syndrome and disease-specific polygenic scores in UK Biobank participants of Asian ancestry and in the ATTRaCT cohort (**Fig. S4**). The comparison included two MetS PGSs (Lind2019_MetS [32] and Walree2022_MetS [33]) and six disease-specific PGSs for type 2 diabetes (Suzuki2024_T2D [34]), heart failure (Levin2022_HF [14]), stroke (Mishra2022_Stroke [35]), myocardial infarction (Hartiala2021_MI [36]), hypertension (Sinnott2021_HTN [37]), and coronary artery disease (Aragam2022_CAD [17]), detailed information about the related GWAS information was reported in **Table S13**.

Polygenic scores were constructed using PRSice2 [38] based on publicly available GWAS summary statistics. For each trait, optimal p-value thresholds were selected to maximise predictive performance in the target dataset. Individual-level genotype data from UK Biobank Asian participants and from the ATTRaCT cohort were used to generate standardized polygenic scores.

Associations between each genetic score and cardiometabolic disease outcomes were estimated using logistic regression models. For each disease, age-balanced case–control datasets were constructed separately within each cohort. Genetic scores were standardised to unit variance, and odds ratios (ORs) with 95% confidence intervals were estimated per one standard deviation increase in each score.

To ensure comparability across predictors and cohorts, association estimates were combined using fixed-effect inverse-variance weighted meta-analysis across the UK Biobank Asian and ATTRaCT cohorts. Meta-analyses were performed separately in women and men, consistent with the sex-stratified analyses used for GenMetS. Across both cohorts, GenMetS showed consistently stronger or comparable associations with type 2 diabetes, heart failure, myocardial infarction, stroke, hypertension, and coronary artery disease relative to previously published metabolic syndrome polygenic scores, and broadly comparable associations to several disease-specific polygenic scores (**Fig. S4**).

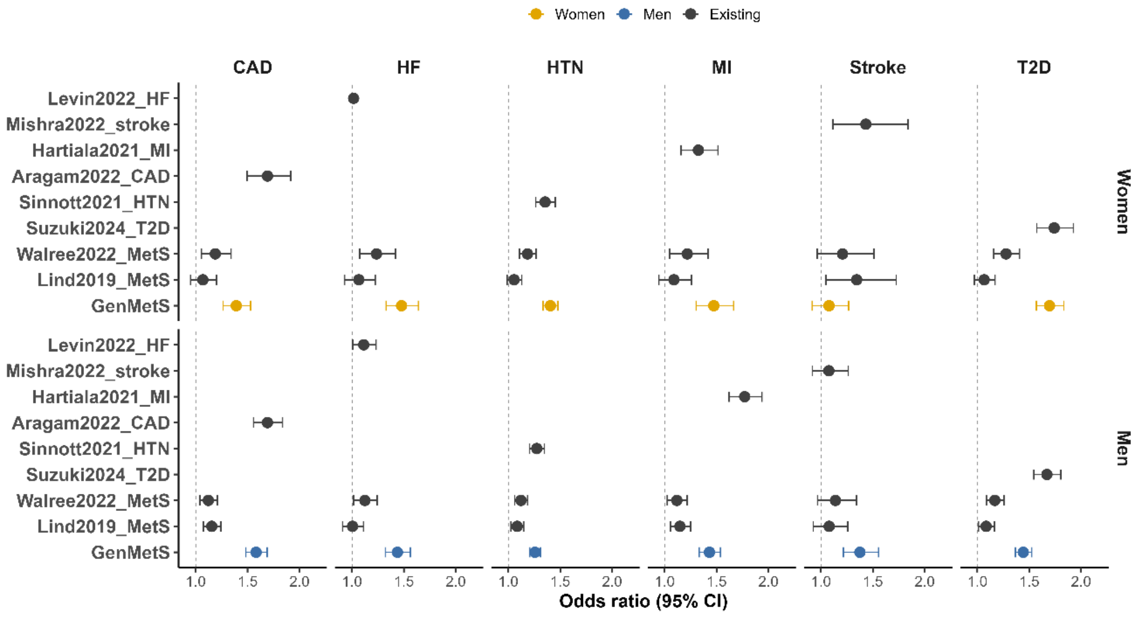


**Fig. S4. Benchmarking GenMetS against published polygenic scores across cardiometabolic diseases and MetS.** Forest plots show odds ratios (ORs) and 95% confidence intervals per standard deviation increase in each score for six cardiometabolic outcomes, stratified by sex. GenMetS was benchmarked against previously published polygenic scores, including metabolic syndrome scores (Lind2019_MetS and Walree2022_MetS) and disease-specific scores where available. Estimates were derived from fixed-effect meta-analyses of UK Biobank participants of Asian ancestry and the ATTRaCT cohort to ensure comparability across predictors. Empty cells indicate that a disease-specific score was not available for the corresponding outcome. GenMetS is highlighted in colour, whereas previously published scores are shown in grey.

**Benchmarking against single-trait polygenic scores in prediction of cardiometabolic diseases**

To benchmark the predictive efficacy of GenMetS, we generated 15 polygenic scores (PGSs) for individual metabolic traits from the resource data (**Table S6**). We studied cardiometabolic diseases, including T2D, CAD, HF, HTN, stroke, and MI in UKB Asian and ATTRacT cohorts.

In the UKB Asian, cases and controls were matched by age and education, ensuring controls were free from the six studied diseases. For ATTRaCT subjects, cases and controls were matched by age without excluding other conditions for the control subjects, given ATTRaCT is a disease cohort.

Benchmarking analysis with the 15 single trait PGS in UKB Asian and ATTRaCT demonstrated the superior predictive power of GenMetS over the majority of single-trait PGSs across multiple cardiometabolic diseases in both genders, **Figs. S5-S6**. It is noteworthy that specific PGSs, particularly those associated with glucose (PGS000306 [29] and PGS000684 [37]) and HbA1c (PGS000685 [37]) demonstrated moderate prediction abilities for T2D. Additionally PGSs related to blood pressure, such as PGS000301 [29] and PGS001134 [24] were predictive for HTN in both UKB Asian and ATTRaCT, with comparable results with GenMetS. Notably, a blood pressure PGS (PGS001133 [24]) outperformed GenMetS in predicting HTN within ATTRaCT cohort.

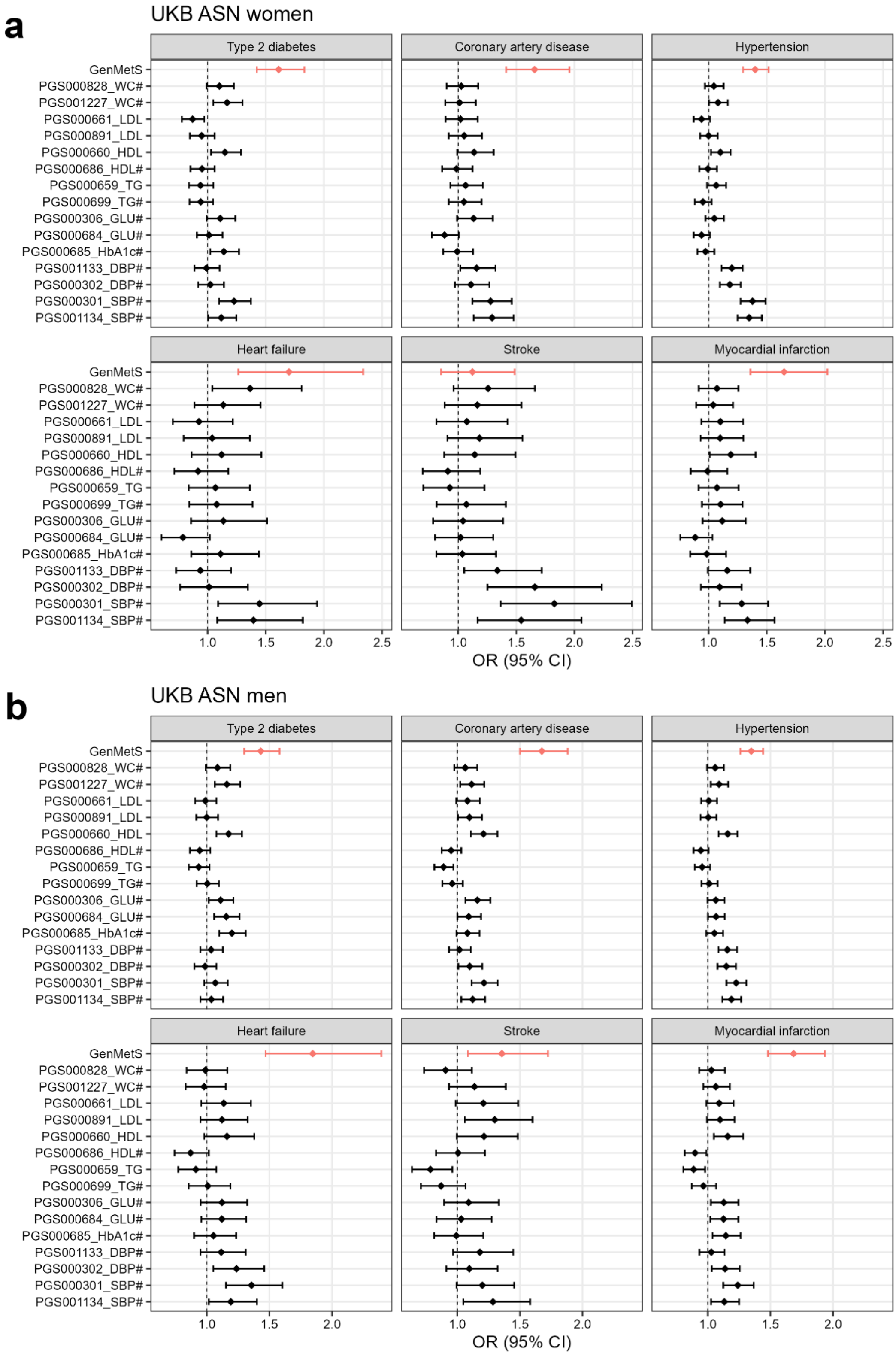


**Fig. S5: GenMetS and other PGSs from MetS traits in prediction of cardiometabolic diseases in UKB Asian (ASN) women and men. a,** UKB Asian women (N= 4,189), **b,** UKB Asian men (N=4,603). Case and control subjects were selected for similar distributions of age and education backgrounds in UKB Asian men. "#" indicates that the polygenic score was learned from cohorts of European ancestry.

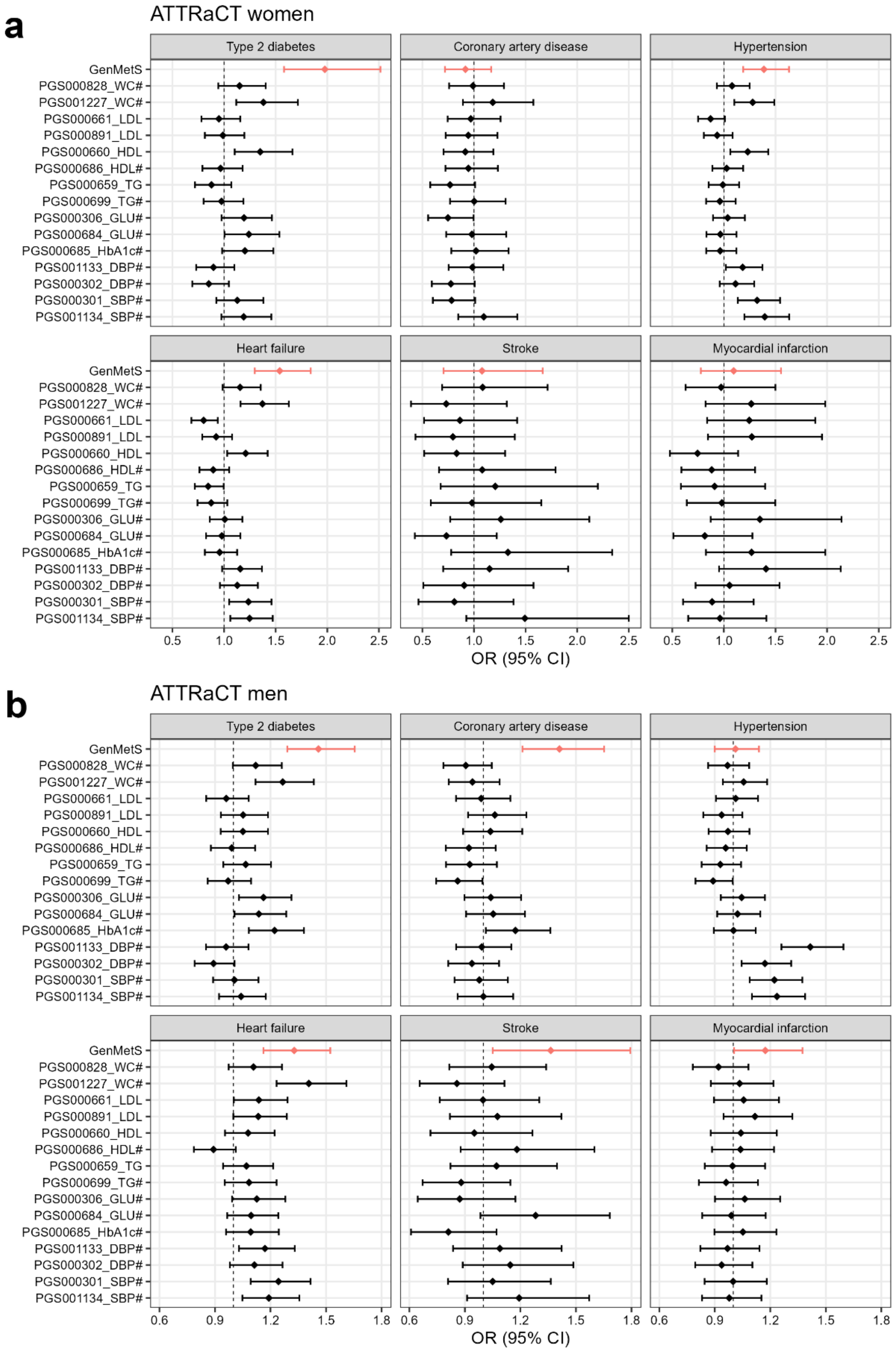


**Fig. S6. GenMetS and other PGSs from MetS traits in prediction of cardiometabolic diseases in ATTRaCT women and men. a,** ATTRaCT women (N= 820), **b,** ATTRaCT men (N=1,490). Case and control subjects were selected for similar distributions of age. “#” indicates that the polygenic score was learned from cohorts of European ancestry.

**Acknowledgement**

**Extreme thanks go to S-PRESTO study group members:**

Airu Chia, Andrea Cremaschi, Anna Magdalena Fogel, Anne Eng Neo Goh, Anne Rifkin-Graboi, Anqi Qiu, Bee Wah Lee, Bernard Su Min Chern, Candida Vaz, Chan Shi Yu, Dawn Xin Ping Koh, Dennis Wang, Desiree Y. Phua, Elaine Phaik Ling Quah, Elizabeth Huiwen Tham, Evelyn Chung Ning Law, Evelyn Keet Wai Lau, Evelyn Xiu Ling Loo, Fabian Kok Peng Yap, Falk Müller-Riemenschneider, George Seow Heong Yeo, Gerard Chung Siew Keong, Hannah Ee Juen Yong, Helen Yu Chen, Hong Pan, Huang Jian, Huang Pei, Hugo P S van Bever, Hui Min Tan, Ives Lim Yubin, Jadegoud Yaligar, Jerry Kok Yen Chan, Jia Xu, Johan Gunnar Eriksson, Jonathan Tze Liang Choo, Jonathan Y. Bernard, Jonathan Yinhao Huang, Jun Shi Lai, Karen Mei Ling Tan, Keith M. Godfrey, Keri McCrickerd, Kok Hian Tan, Kok Wee Chong, Kothandaraman Narasimhan, Kuan Jin Lee, Li Chen, Lieng Hsi Ling, Ling-Wei Chen, Lourdes Mary Daniel, Lynette Pei-Chi Shek, Maria De Iorio, Marielle V. Fortier, Mary Foong-Fong Chong, Mary Wlodek, Mei Chien Chua, Melvin Khee-Shing Leow, Michael J. Meaney, Michelle Zhi Ling Kee, Min Gong, Mya Thway Tint, Navin Michael, Neerja Karnani, Ngee Lek, Noor Hidayatul Aini Bte Suaini, Oon Hoe Teoh, Peter David Gluckman, Priti Mishra, Queenie Ling Jun Li, Sambasivam Sendhil Velan, See Ling Loy, Seng Bin Ang, Shiao-Yng Chan, Shirong Cai, Shu-E Soh, Si Hui Goh, Stephen Chin-Ying Hsu, Suresh Anand Sadananthan, Tan Ai Peng, Teng Hong Tan, Varsha Gupta, Victor Samuel Rajadurai, Wee Meng Han, Wei Wei Pang, Yap Seng Chong, Yin Bun Cheung, Yiong Huak Chan, Yung Seng Lee, Zai Ru Cheng, Zhang Han

**Growing Up in Singapore Towards healthy Outcomes (GUSTO)**

GUSTO is a mother-offspring prospective cohort study in Singapore [2]. **GUSTO** study is supported by the National Research Foundation (NRF) under the Open Fund-Large Collaborative Grant (OF-LCG; MOH-000504) administered by the Singapore Ministry of Health's National Medical Research Council (NMRC) and the Agency for Science, Technology and Research (A*STAR). In RIE2025, GUSTO is supported by funding from the NRF's Human Health and Potential (HHP) Domain, under the Human Potential Programme.

**Extreme thanks go to GUSTO study group members:**

This study group includes: Airu Chia, Andrea Cremaschi, Anna Magdalena Fogel, Anne Eng Neo Goh, Anne Rifkin-Graboi, Anqi Qiu, Arijit Biswas, Bee Wah Lee, Birit Froukje Philipp Broekman, Candida Vaz, Chai Kiat Chng, Chan Shi Yu, Choon Looi Bong, Daniel Yam Thiam Goh, Dawn Xin Ping Koh, Dennis Wang, Desiree Y. Phua, E Shyong Tai, Elaine Kwang Hsia Tham, Elaine Phaik Ling Quah, Elizabeth Huiwen Tham, Evelyn Chung Ning Law, Evelyn Keet Wai Lau, Evelyn Xiu Ling Loo, Fabian Kok Peng Yap, Falk Müller-Riemenschneider, Franzolini Beatrice, George Seow Heong Yeo, Gerard Chung Siew Keong, Hannah Ee Juen Yong, Helen Yu Chen, Hong Pan, Huang Jian, Huang Pei, Hugo P S van Bever, Hui Min Tan, Iliana Magiati, Inez Bik Yun Wong, Ives Lim Yubin, Ivy Yee-Man Lau, Jacqueline Chin Siew Roong, Jadegoud Yaligar, Jerry Kok Yen Chan, Jia Xu, Johan Gunnar Eriksson, Jonathan Tze Liang Choo, Jonathan Y. Bernard, Jonathan Yinhao Huang, Joshua J. Gooley, Jun Shi Lai, Karen Mei Ling Tan, Keith M. Godfrey, Keri McCrickerd, Kok Hian Tan, Kothandaraman Narasimhan, Krishnamoorthy Naiduvaje, Kuan Jin Lee, Li Chen, Lieng

Hsi Ling, Lin Lin Su, Ling-Wei Chen, Lourdes Mary Daniel, Lynette Pei-Chi Shek, Maria De Iorio, Marielle V. Fortier, Mary Foong-Fong Chong, Mary Wlodek, Mei Chien Chua, Melvin Khee-Shing Leow, Michael J. Meaney, Michelle Zhi Ling Kee, Min Gong, Mya Thway Tint, Navin Michael, Neerja Karnani, Ngee Lek, Noor Hidayatul Aini Bte Suaini, Oon Hoe Teoh, Peter David Gluckman, Priti Mishra, Queenie Ling Jun Li, Sambasivam Sendhil Velan, Seang Mei Saw, See Ling Loy, Seng Bin Ang, Shang Chee Chong, Shiao-Yng Chan, Shirong Cai, Shu-E Soh, Stephen Chin-Ying Hsu, Suresh Anand Sadananthan, Swee Chye Quek, Tan Ai Peng, Varsha Gupta, Victor Samuel Rajadurai, Wee Meng Han, Wei Wei Pang, Yap Seng Chong, Yin Bun Cheung, Yiong Huak Chan, Yung Seng Lee, Zhang Han

**The Asian neTwork for Translational Research and Cardiovascular Trials (ATTRaCT)**

ATTRaCT is supported by the following grants: NMRC grant – CVRI Centre Grant (NMRC/CG/014/2013), NRMC grant - NMRC Translational and Clinical Research (TCR) Flagship Programme Tier 1 (NMRC/TCR/006-NUHS/2013) and the A*STAR BMRC – Strategic Positioning Fund (SPF) grant (Reference number SPF2014/003; SPF2014/004; SPF2014/005) This study was conducted according to the guidelines laid down in the Declaration of Helsinki. All participants provided written informed consent.

**Extreme thanks go to ATTRaCT study group members:**

The study group includes: Jasper Tromp, Wouter Ouwerkerk, Calvin Chin Woon Loong, Vera Goh Jin-Ling, Tong Jieli, Yap Siew Fei, Jin Xuanyi, Sheldon Lee Shao Guang, Soon Dinna, Angela Koh Su Mei, Carolyn Lam, Lieng Hsi Ling, Li San Lynette Teo, Tze Pin Ng, Raymond C Wong, Kar Yin Se, Ping Chai, Kian Keong Poh, Mark Richards, Seet Yoong Loh, Poh Shuan Daniel Yeo, Lee Fong Ling, Hean Yee Ong, Yong Quek Wei Yong, Changfen Xu, Jaufeerally Fazlur Rehman, Patrick Cozzone, Han Weiping, Philip Le, Stuart Cook, Tan Ru San, Angela Koh, David Townsend, John Totman, Peter Little, Dominique de Kleijn, Colin Stewart, Shaun Loong, Roger Foo, Ng Huck Hui, JJ Liu, Mark Richards, Anis Larbi, Brian Abel, Alessandra Nardin, Michael Poidinger, Laurent Renia, Jayatha Gunaratne, Salvatore Albani, Theodoros Kofidis

**Biobank Japan (BBJ)**

The BioBank Japan (BBJ) is a hospital-based national biobank project that collects DNA and serum samples and clinical information from 12 medical institutes in Japan. BBJ is supported by the Tailor-Made Medical Treatment Program of the Ministry of Education, Culture, Sports, Science, and Technology (MEXT) and AMED under grant numbers JP17km0305002 and JP17km0305001, JP.24tm0624002.

**Extreme thanks go to BBJ Project Consortium members:**

This study group includes Koichi Matsuda, Yukinori Okada, Yuji Yamanashi, Yoichi Furukawa, Takayuki Morisaki, Yoshinori Murakami, Yoichiro Kamatani, Kaori Muto, Akiko Nagai, Wataru Obara, Ken Yamaji, Kazuhisa Takahashi, Satoshi Asai, Yasuo Takahashi, Takao Suzuki, Nobuaki Sinozaki, Hiroki Yamaguchi, Shiro Minami, Shigeo Murayama, Kozo Yoshimori, Satoshi Nagayama, Daisuke Obata, Masahiko Higashiyama, Akihide Masumoto, Yukihiro Koretsune, Yukihide Momozawa, Chikashi Terao.

**Members of the China Kadoorie Biobank collaborative group:**

**International Steering Committee:** Junshi Chen, Zhengming Chen (PI), Robert Clarke, Rory Collins, Liming Li (PI), Jun Lv, Richard Peto, Robin Walters.

**International Co-ordinating Centre, Oxford:** Daniel Avery, Maxim Barnard, Derrick Bennett, Ruth Boxall, Ka Hung Chan, Yiping Chen, Zhengming Chen, Charlotte Clarke, Johnathan Clarke; Robert Clarke, Huaidong Du, Geoffrey Ma, Ahmed Edris Mohamed, Hannah Fry, Simon Gilbert, Pek Kei Im, Andri Iona, Maria Kakkoura, Christiana Kartsonaki, Kshitij Kolhe, Hubert Lam, Kuang Lin, James Liu, Mohsen Mazidi, Iona Millwood, Sam Morris, Qunhua Nie, Alfred Pozarickij, Maryam Rahmati, Paul Ryder, Dan Schmidt, Becky Stevens, Iain Turnbull, Robin Walters, Baihan Wang, Lin Wang, Neil Wright, Ling Yang, Xiaoming Yang, Pang Yao.

**National Co-ordinating Centre, Beijing:** Xiao Han, Can Hou, Qingmei Xia, Chao Liu, Jun Lv, Pei Pei, Dianjianyi Sun, Canqing Yu, Lang Pan.

**10 Regional Co-ordinating Centres:**

**Qingdao CDC:** Zengchang Pang, Ruqin Gao, Shanpeng Li, Haiping Duan, Shaojie Wang, Yongmei Liu, Ranran Du, Yajing Zang, Liang Cheng, Xiaocao Tian, Hua Zhang, Yaoming Zhai, Feng Ning, Xiaohui Sun, Feifei Li. **Licang CDC:** Silu Lv, Junzheng Wang, Wei Hou. **Heilongjiang Provincial CDC:** Wei Sun, Shichun Yan, Xiaoming Cui. **Nangang CDC:** Chi Wang, Zhenyuan Wu,Yanjie Li, Quan Kang. **Hainan Provincial CDC:** Huiming Luo, Tingting Ou. **Meilan CDC:** Xiangyang Zheng, Zhendong Guo, Shukuan Wu, Yilei Li, Huimei Li. **Jiangsu Provincial CDC:** Ming Wu, Yonglin Zhou, Jinyi Zhou, Ran Tao, Jie Yang, Jian Su. **Suzhou CDC:** Fang Liu, Jun Zhang, Yihe Hu, Yan Lu, Liangcai Ma, Aiyu Tang, Shuo Zhang, Jianrong Jin, Jingchao Liu. **Guangxi Provincial CDC:** Mei Lin, Zhenzhen Lu. **Liuzhou CDC:** Lifang Zhou, Changping Xie, Jian Lan,Tingping Zhu,Yun Liu, Liuping Wei, Liyuan Zhou, Ningyu Chen, Yulu Qin, Sisi Wang. **Sichuan Provincial CDC:** Xianping Wu, Ningmei Zhang, Xiaofang Chen, Xiaoyu Chang. **Pengzhou CDC:** Mingqiang Yuan, Xia Wu, Xiaofang Chen, Wei Jiang, Jiaqiu Liu, Qiang Sun. **Gansu Provincial CDC:** Faqing Chen, Xiaolan Ren, Caixia Dong. **Maiji CDC:** Hui Zhang, Enke Mao, Xiaoping Wang, Tao Wang, Xi zhang. **Henan Provincial CDC:** Kai Kang, Shixian Feng, Huizi Tian, Lei Fan. **Huixian CDC:** XiaoLin Li, Huarong Sun, Pan He, Xukui Zhang. **Zhejiang Provincial CDC:** Min Yu, Ruying Hu, Hao Wang. **Tongxiang CDC**: Xiaoyi Zhang, Yuan Cao, Kaixu Xie, Lingli Chen, Dun Shen. **Hunan Provincial CDC:** Xiaojun Li, Donghui Jin, Li Yin, Huilin Liu, Zhongxi Fu. **Liuyang CDC:** Xin Xu, Hao Zhang, Jianwei Chen,Yuan Peng, Libo Zhang, Chan Qu.

**Funding:**

The CKB baseline survey and the first re-survey were supported by the Kadoorie Charitable Foundation in Hong Kong. Long-term follow-up was supported by Wellcome grants to Oxford University (212946/Z/18/Z, 202922/Z/16/Z, 104085/Z/14/Z, 088158/Z/09/Z) and grants from the National Key Research and Development Program of China (2016YFC0900500, 2016YFC0900501, 2016YFC0900504, 2016YFC1303904) and the National Natural Science Foundation of China (91843302). DNA extraction and genotyping was supported by grants from GlaxoSmithKline and the UK Medical Research Council (MC-PC- 13049, MC-PC-14135). The UK Medical Research Council (MC_UU_00017/1,MC_UU_12026/2 MC_U137686851), Cancer Research UK (C16077/A29186; C500/A16896) and the British Heart Foundation (CH/1996001/9454) provided core funding to the Clinical Trial Service Unit and Epidemiological Studies Unit at Oxford University for the project. Computation used the Oxford Biomedical Research Computing (BMRC) facility, a joint development between the Wellcome Centre for Human Genetics and the Big Data Institute supported by Health Data Research UK and the NIHR Oxford Biomedical Research Centre; the views expressed are those of the author(s) and not necessarily those of the NHS, the NIHR or the Department of Health.
